# SARS-CoV-2 Genetic Variants and Patient Factors Associated with Hospitalization Risk

**DOI:** 10.1101/2024.03.08.24303818

**Authors:** Tonia Korves, David Stein, David Walburger, Tomasz Adamusiak, Seth Roberts

**Author notes:** Corresponding author: Seth Roberts,. Center for Clinical investigation, Brigham and Women’s Hospital, 221 Longwood Ave, Boston, Massachusetts 02115 (David Stein).

## Abstract

Variants of SARS-CoV-2 have been associated with different transmissibilities and disease severities. The present study examines SARS-CoV-2 genetic variants and their relationship to risk for hospitalization, using data from 12,538 patients from a large, multisite observational cohort study. The association of viral genomic variants and hospitalization is examined with clinical covariates, including COVID-19 vaccination status, outpatient monoclonal antibody treatment status, and underlying risk for poor clinical outcome. Modeling approaches include XGBoost with SHapley Additive exPlanations (SHAP) analysis and generalized linear mixed models. The results indicate that several SARS-CoV-2 lineages are associated with increased hospitalization risk, including B.1.1.7, AY.44, and AY.54. As found in prior studies, Omicron is associated with lower hospitalization risk compared to prior WHO variants. In addition, the results suggest that variants at specific amino acid locations, including locations within Spike protein N-terminal domain and in non-structural protein 14, are associated with hospitalization risk.

## INTRODUCTION

Since the start of the COVID-19 pandemic, SARS-CoV-2 lineages have evolved to differ in transmissibility, disease severity, interactions with the human immune system, and susceptibility to vaccines (1). Understanding how SARS-CoV-2 genetic variants impact health outcomes is important for surveillance and the future development of therapeutics and vaccines.

Multiple studies have investigated associations between SARS-CoV-2 genomic variation and COVID severity in different times and places during the pandemic and collectively have identified a diverse set of candidate genetic variants (2–16). These studies have either used data from GISAID (2, 4–6, 9, 11, 16–18) or from individual hospitals (3, 10, 12–15). The GISAID-based studies use COVID-19 severity metrics that classify samples into two or more severity categories, such as ‘asymptomatic’ and ‘severe’, based on patient status descriptions. These studies typically use datasets from a few thousand to more than ten thousand samples. Most of these studies include patient age and sex as covariates in analyses, but, because GISAID lacks additional patient data, they do not include patient co- morbidity information, COVID-19 vaccination status, or drug treatments. These factors are known to have substantial consequences for COVID health outcomes and absence of data about them may obscure genetic associations. Genome-wide association studies (GWAS) using data from individual hospitals have evaluated differences in outcome severity among hospitalized patients. These studies often incorporate extensive co-morbidity and other patient data, but typically have only hundreds of samples.

In addition to data availability limitations, investigating associations between SAR-CoV-2 genetic variants and health outcomes is challenging due to the virus’s largely clonal structure. Single nucleotide variants are often tightly correlated with each other, and these correlations need to be accounted for to disambiguate associations. Some studies have used methods to analyze one variant at a time, including chi-square or Fisher exact tests that do not account for genetic correlations (2, 3), multiple logistic regression with covariates to capture genetic background like in many human GWAS (4, 14, 15), and phylogeny-based association tests (6). Other studies have used analytical approaches to investigate multiple variant loci at a time, including multiple logistic regression with multiple variants as features (5, 17), neural network approaches (7, 8), random forests (16, 17), and XGBoost (7, 8).

XGBoost offers several advantages for investigating SARS-CoV-2 variant-health outcome associations. It has been a top-performing algorithm for many classification problems, can incorporate many features at once, enables the investigation of interactions between features, is fairly robust to collinearity, and has ways for handling imbalanced outcome data (19, 20). XGBoost can be used with Shapley Additive exPlanations (SHAP) analysis, an explainable artificial intelligence method based on game theory that measures the contribution each feature makes to a model’s predictions (21). Another method that has been used recently to study SARS-CoV-2 genetic variant fitness is hierarchical Bayesian multinomial logistic regression; this method provides a way to account for lineage associations and to calculate the uncertainty of parameter estimates (22).

In this paper, we present an analysis of SARS-CoV-2 genomic variants and clinical outcomes using a large dataset from a study on the effectiveness of neutralizing monoclonal antibodies (nMAbs) for SARS-CoV-2 (23, 24). The nMAbs observational study collected data from four health care systems in the United States from November 2020 through January 2022 and included patients diagnosed with COVID-19 who met at least one Emergency Use Authorization “high risk” criterion supporting the use of nMAbs (25). The study demonstrated an association between nMAbs treatment and reduced odds of hospitalization and death, with larger treatment effects in unvaccinated patients and those with higher risk of poor outcomes (23). For 13,703 of the patients in the study, SARS-CoV-2 diagnostic samples were collected and sequenced. This genomic data was used to identify WHO variants, Pango lineages, and amino acid mutations that may confer an escape capability to nMab treatments (24).

In this study, we used the subset of nMAbs observational study patients with both clinical data and viral genomic data to identify candidate SARS-CoV-2 genetic variants associated with altered odds of hospitalization within 14 days of diagnosis. We used two approaches, 1) XGBoost modeling and SHAP analysis to investigate associations across the whole dataset and 2) generalized linear mixed models to investigate associations between and within SARS-CoV-2 lineages. The analyses accounted for each patient’s underlying disease risk due to demographics and comorbidities, nMAb treatment, and COVID-19 vaccination status, and explored interactions between these clinical covariates and genetic variants. We found evidence that risk for hospitalization varies among SARS-CoV-2 lineages and among variants at specific amino acid locations, including locations within spike (S) protein and non-structural protein 14 (nsp14).

## METHODS

The data used for this study come from a prior observational cohort study of the effectiveness of nMAbs. Details of this dataset have been described previously (23) and are described in Supplementary Materials. In brief, the nMAbs dataset includes structured clinical data (e.g., demographic features, comorbidities, COVID-19 vaccination and nMAb treatment status, outcomes) and linked viral genomic data (e.g., complete SARS-CoV-2 genome sequences) for 13,703 patients from four health systems within the United States. The viral genomic data were derived from patient samples taken on the day of COVID-19 diagnosis.

### Patient Covariates

For each patient, we derived a Boolean feature, nMAb treatment status, that represents whether or not the patient was treated with an nMAb for COVID-19. Following the parent observational cohort study, a patient was considered “treated” if they received any of the four nMAbs treatments available during the study period within 10 days of their diagnosis.

We also derived a second Boolean feature, COVID-19 vaccination status, that represents whether or not the patient was vaccinated to some degree (including partial or incomplete primary course, completed primary course, or completed primary course with one or more booster doses). Patients with no evidence of any COVID-19 vaccination were considered “unvaccinated”.

We developed a predictive model to summarize each patient’s “underlying risk” for poor COVID-19 outcomes, given their specific demographics and combination of comorbidities. We used the model to encapsulate each patient’s risk due to these factors in a single number, the disease risk score. To create the model, we included the following patient features as independent variables: age decile (10-year groupings), gender, comorbidities grouped according to Elixhauser (26), ethnicity, race, marital status, insurance type, total prior healthcare visits, population density and area deprivation index (28) for the zip3 of patient primary residence, and indicator variables for smoking status, pregnancy, obesity, and whether a patient had recently taken immunosuppressant medication. Categorical features with more than one level were one-hot encoded. These included age group, gender, ethnicity, race, marital status, insurance type, and smoking status. Continuous features, including zip3 population density and area deprivation index, and total prior healthcare visits, were scaled into the [0,1] interval. The list of features used in disease risk score modeling is in the Supplementary Materials, Table S1. Of note, among the features not used in disease risk modeling were COVID-19 vaccination status and nMAb treatment status, as we wanted to explicitly represent any association of these with clinical outcomes in further modeling. In addition, date of infection, which is associated with SARS-CoV-2 genetic variant, was not included as a feature for disease risk score given the goal of assessing genetic associations.

This set of features was used to predict whether death or hospitalization occurred within 30 days of the date of diagnosis, among all patients who were not treated with nMAbs, including patients with and without SARS-CoV-2 genomic data. A training set was constructed using a random 90% subset of the untreated patients (141,942 total untreated patients, of which 127,748 used for the training set), with the remaining 10% held out for testing (n=14,194). The scaling of continuous variables was derived using the training dataset and applied to the test set. For modeling, we used the scikit-learn Python package (29). Specifically, we used `sklearn.linear_model.LogisticRegression` with the following hyperparameters: l2 penalty, inverse of regularization strength (C) of 10, balanced class weighting, maximum iterations of 1000, and the ‘lbfgs’ solver. After fitting the model on the training set, the model was evaluated on the test set using area under the receiver operating characteristic curve (AUROC) and area under the precision-recall curve (PR-AUC). Following model evaluation, a disease risk score was calculated for all persons in the dataset. The score represents an estimated probability of death or hospitalization within 30 days in the absence of nMAb treatment, given the input features.

### Identification of Lineages, WHO Variants, and Amino Acid Changes

Details regarding our genomics data processing pipeline can be found in Supplementary Materials. Briefly, the pipeline processed FASTQ file inputs and generated consensus fasta genome (DNA) sequences for all samples; 12,538 samples met data quality thresholds and were used in analyses. To address missing base calls, the pipeline included a step for imputation of missing or ambiguous sequence data using UShER (30); we term the resulting consensus sequences UShER-cleaned sequences (see Supplementary Materials).

We processed the UShER-cleaned sequences via the pangolin tool for implementing the dynamic nomenclature of SARS-CoV-2 lineages (31), version 4.2 with pangolin-data version 1.19, to produce updated SARS-CoV-2 lineage calls. We then used the lineage designations to assign sequences to WHO variants (Supplemental Table S2). To identify variants at the amino acid level, the UShER-cleaned DNA sequences were processed with the Nextclade tool for mutation calling and sequence quality checks (32). Amino acid variant data were condensed to represent all alternate substitutions at a single protein-amino acid position as one feature (i.e., 1 for any alternate variant, 0 for the Wuhan reference strain variant); stop codon amino acid positions were kept as distinct features. We refer to these features as “amino acid changes”.

For the association analyses, we selected a subset of amino acid changes to evaluate. First, to mitigate collinearity, we eliminated amino acid changes that were highly correlated with other amino acid changes characteristic of a particular WHO variant. To do this, we eliminated amino acid changes that were correlated with one of the six common WHO variants (Alpha, Delta, Epsilon, Gamma, Mu, or Omicron) at greater than 0.9, and instead used WHO variant as a feature to collectively account for these sets of amino acid changes. Second, we applied a minimum allele frequency (MAF) and assessed only amino acid changes that occurred with a frequency between 0.01 and 0.99 within the dataset. Finally, to prevent the inclusion of artifacts from the sequence imputation strategy described in Supplementary Materials, we removed all amino acid changes where the majority of samples with the alternate allele did not have an alternate allele in the original data before imputation.

### XGBoost and SHAP analyses

To evaluate associations between hospitalization within 14 days and genetic variation across lineages, we trained XGBoost models with the following features: amino acid changes, WHO variant designations, and the three clinical patient covariates (disease risk score, COVID-19 vaccination status, and nMAb treatment status). WHO variant was one-hot encoded, with one feature for each of the six most common WHO variants in the dataset.

To ensure robust performance and to calculate SHAP values across the entire dataset, we used repeated k-fold cross-validation, with 5-fold cross validation with 80% training / 20% cross-validation data, repeated for five different randomly selected ways of splitting the data, resulting in a set of 25 data split-folds. For each fold from each split, we used the training data to identify hyperparameters by performing a random grid search using `sklearn.model_selection.RandomizedSearchCV` (29) (see Supplementary Materials Table S3 for hyperparameter search grid values). The assessment metric used in hyperparameter selection was `average_precision` which measures area under the precision-recall curve (PR-AUC). To adjust for imbalance in the outcome variable, in all models, we used a scale- weight value equal to the number not hospitalized in the data divided by number hospitalized. XGBoost models were trained with the R xgboost package (33) using the hyperparameter values identified for the corresponding data split-fold training data. To account for random variation in model construction, for each of the 25 data split-folds, five XGBoost models were constructed with different seed initializations, and the median result values, including median performance metric and SHAP values, were selected for each data split-fold for further analyses.

Because we were primarily interested in predicting cases of hospitalization, which were infrequent (3-4% of cases), we evaluated the models’ predictive ability by calculating PR-AUC. For each data split-fold, we used the cross-validation set to calculate PR-AUC with the package MLmetrics (34). To assess whether the genetic variation features contributed to predictive ability, XGBoost models were also created using just the three clinical features, excluding all amino acid changes and WHO variant features. This was done with the same folds and splits of the data as used for the model with the full set of features, and the same grid search procedure for selection of hyperparameters. PR-AUC values were compared between models with all features and models with just the three clinical covariates by doing a one-sided Wilcoxon test using paired results from each of the 25 data split folds.

To identify important features, we performed SHAP analysis (35). SHAP values and SHAP interaction values were calculated on the training data for each of the data split-folds using the R xgboost package (33). For each of the 25 data split-folds, we calculated mean absolute SHAP value for each feature across samples and mean absolute SHAP interaction values between pairs of features across all samples. To identify important features, we calculated the means of these across all the data split-folds, and features were sorted by overall mean absolute SHAP value, from highest to lowest. For top-ranked features, we calculated descriptive summary statistics about their variation and calculated median SHAP values for each sample across the 25 data split-folds. These values were used to create SHAP beeswarm plots using the R shapviz package (36) and generate plots using ggplot2 (37) to examine relationships between SHAP values and features.

### Generalized linear mixed models

For analysis with generalized linear mixed models (GLMMs), we chose a subset of the amino acid changes studied in XGBoost and SHAP analyses, because the GLMMs were designed to assess associations of amino acid changes within lineages. Thus, the amino acid changes chosen had the following characteristics within at least one Pango lineage: amino acid change represented by at least 50 samples and frequency < 0.75. We set a minimum of 50 samples to avoid testing extremely rare amino acid changes. We enforced a frequency of < 0.75 in the same lineage to avoid testing amino acid changes highly collinear with lineage, as lineage-level effects are explicitly included in the GLMMs.

To assess for potential associations between amino acid changes at the sub-lineage level and hospitalization, we used GLMMs and Bayesian techniques for parameter estimation. In this approach, risk of hospitalization within 14 days is predicted using clinical variables (nMAb treatment status, COVID-19 vaccination status, disease risk score), lineage-level effects (separate intercepts for each lineage), and the effect of a single amino acid change, where the effect may be different across different lineages. Parameters were estimated via stochastic variational inference (38) using Pyro (39, 40), a probabilistic programming language for Python. We ran this model separately for each of the amino acid changes identified for analysis at the sub-lineage level (as described below, 28 amino acid changes were identified for analysis with GLMMs; thus, we ran this model 28 times, once for each amino acid change in turn). After specifying the model and running stochastic variational inference, the trained model was used to produce posterior distributions for all model parameters, including lineage intercept effect estimates and lineage-specific amino acid change effect estimates. Our model can be expressed as:

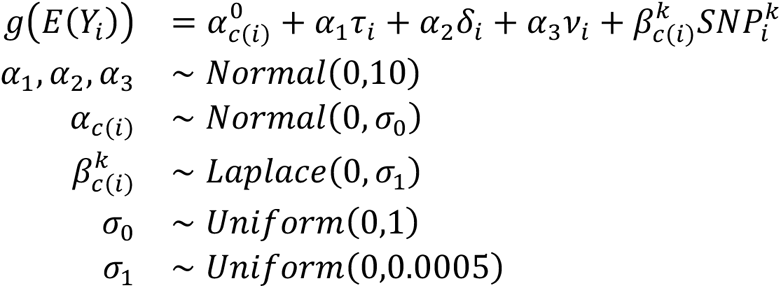

where *Y_i_* is the outcome of patient *i*; *g* is the logit link function; *α*_c(*i*)_ represents the intercept for lineage *c*(*i*); *τ*, *δ*, and *v* denote patient nMAb treatment status, disease risk score, and COVID-19 vaccination status, respectively; *SNP_i_^k^* represents the presence or absence of a specific amino acid change; *Normal*(*x*, *y*) represents a normal prior with location *x* and scale *y*; *Uniform* (*a*, *b*) represents a uniform prior bounded between *a* and *b*; and *Laplace x*, *y*) represents a Laplace prior with location *x* and scale *y*. The intercept *α*_c(*i*)_ and the amino acid change parameter *β^k^_c_*(*i*) are lineage dependent random effects. The scales of these random effects, *σ*_0_ and *σ*_1_, are independent of the lineage. The nMAb treatment, disease risk score, and vaccine effects (*α*_1_, *α*_2_, and *α*_3_) are fixed effects. The model is depicted diagrammatically in Figure 1. Broad priors were chosen for nMAb treatment status, disease risk score, and COVID-19 vaccination status (scale = 10). A narrower prior was chosen for the lineage intercept estimates (scale = 1) and, following Obermeyer et al. (22), a very narrow prior was chosen for individual amino acid change effects (scale = 0.0005). Additional analyses were conducted with different priors and are presented in Supplementary Materials.

**Figure 1.**
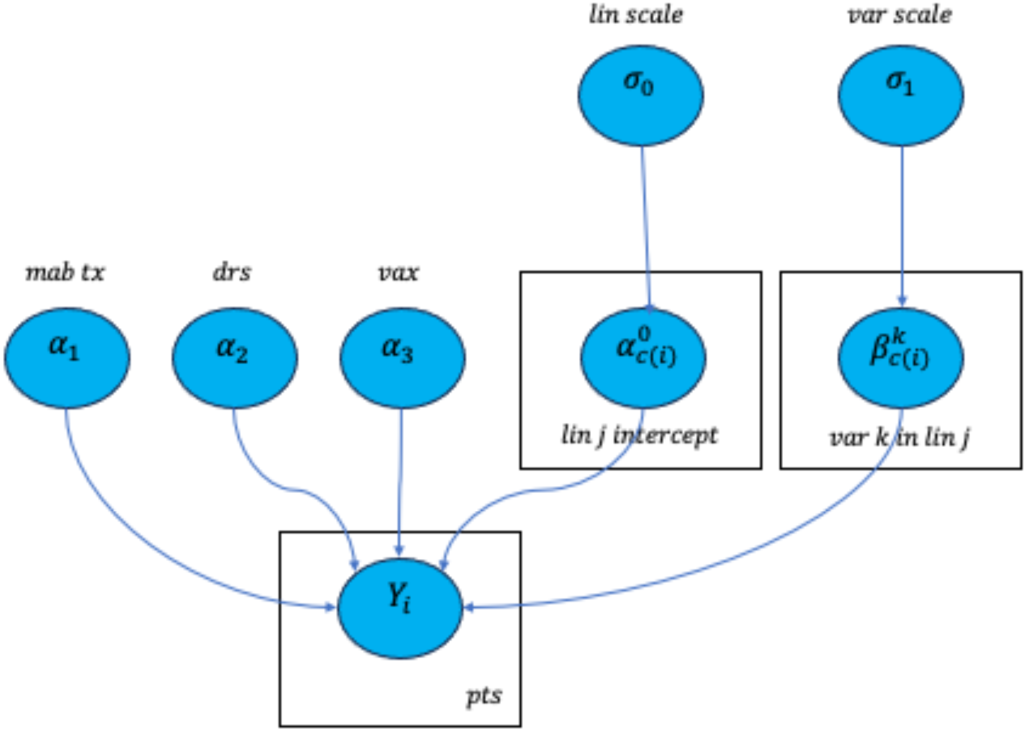
Generalized linear mixed model. The diagram depicts parameters for nMAb treatment (mab tx, *α*_1_), disease risk score (*α*_2_), COVID-19 vaccination status (vax, *α*_3_), shared scale for lineage intercepts (lin scale, *σ*_0_), shared scale for amino acid change across lineages (var scale, *σ*_1_), lineage intercepts (`lin j intercept`, *α*^0^_*c*(*i*)_), and amino acid change for different lineages (`var k in lin j`, *β*^*k*^_*c*(*i*)_. Rectangles represent plates, which are stand- ins for many similar types of variables. For example, the rectangle around *α*^0^_*c*(*i*)_ represents the intercept estimates for all 160 lineages, individually indicated as `c(i)`. The oval *Y_i_* indicates the observed hospitalization outcomes for each patient.

To highlight amino acid changes with the strongest evidence of association with the outcome, we selected amino acid changes with results meeting either of the following two criteria. First, we used the mean and standard deviation of the posterior distributions to calculate z-scores and selected amino acid changes with extreme z-scores (> 1.6 or < -1.6, outside the 90% confidence level centered on zero). Second, we calculated Bayes factors (41) for effect estimates of all amino acid changes and selected those with Bayes factors > 3 (42).

### Visualization of amino acid changes within SARS-CoV-2 proteins

The locations of amino acid changes within SARS-CoV-2 proteins were visualized using x-ray crystallography models from the PDB Database (43) where available and using models predicted with AlphaFold2 (44) where not available. For models predicted with AlphaFold2, amino acid variant substitutions were made in ChimeriaX (45) using the most common amino acid rotamer for the amino acid and surrounding amino acid neighbors. Post-substitution, models were relaxed using SCWRL4 (46) and revisualized in ChimeraX.

## RESULTS

### Clinical and viral genetic variation in the dataset

After filtering for sequence quality, the data included 12,538 patients with genomic sequences (see Supplemental Text). In this set, 3.9% of patients experienced a hospitalization within 14 days of COVID-19 diagnosis. Forty-two percent of patients had a record of COVID-19 vaccination prior to COVID-19 diagnosis, and 33% were treated with nMAbs. Disease risk scores were calculated for these patients using the model trained on demographic and clinical data (this model had a PR-AUC of 0.12 on the test set, see Supplemental Text).

A total of 160 distinct lineages were found across the samples (Table S2). Delta was the most prevalent WHO variant (62% of samples), followed by Omicron (19%), Alpha (9%), Epsilon (2%), Mu (0.6%) and Gamma (0.5%). In addition, 6% of samples, which mostly pre- dated Alpha, were not assigned to a WHO variant, and 0.2% of samples were assigned to other WHO variants.

Across the dataset, 5,189 amino acid changes were identified, which consisted of 5,045 sites with an alternate amino acid substitution compared to the Wuhan reference sequence and 144 sites with stop codons. Of these, 77 were highly correlated (Person *r*^2^ > 0.9) with one of the six most prevalent WHO variants in the dataset; these amino acid changes were removed and instead represented by features for the respective WHO variants (e.g., Omicron, Delta, Alpha). Of the remaining amino acid changes, 4,995 had a minor allele frequency less than 0.01 and were not included in subsequent analyses. An additional 35 amino acid changes were removed to eliminate potential artifacts related to the UShER-based imputation strategy. The remaining 82 amino acid variant sites were used as features in XGBoost modeling. Of these, a subset of 28 amino acid changes that were variable within at least one lineage were used in generalized linear mixed models (GLMMs). Amino acid changes that were evaluated and those that were associated with WHO variants are listed in the Supplemental Materials.

### XGBoost and SHAP analysis results

XGBoost models with clinical, WHO variant, and amino acid change features predicted hospitalization within 14 days with a mean PR-AUC of 0.115. This exceeds the PR-AUC values for the chance, baseline model, which is the frequency of hospitalization in this dataset, 0.039.

XGBoost models with all the features predicted hospitalization better than models built with just the three clinical covariates, indicating that the genetic features contributed to prediction. PR-AUC values for models with all the features were larger than the PR-AUC values for models with just the clinical features across the 25 data split-folds (one-sided, paired Wilcoxon signed-rank test, p=2.3 X10-5). The models with just the clinical covariates had a mean PR-AUC value of 0.103, which is smaller than the mean for the models with all the features. Together, these results indicated that the genetic features collectively made a small but significant contribution to the prediction of hospitalization.

Consistent with this, in SHAP analyses of the models with all the features, the three clinical covariates had the largest contributions to the hospitalization predictions. Table 1 shows the features with the highest mean absolute SHAP values; each value indicates the average magnitude of the effect of a feature on predicted hospitalization risk across all samples. Figure 2 shows that higher disease risk scores predicted that patients were more likely to be hospitalized, and that COVID-19 vaccination and nMAb treatment predicted that patients were less likely to be hospitalized.

**Figure 2.**
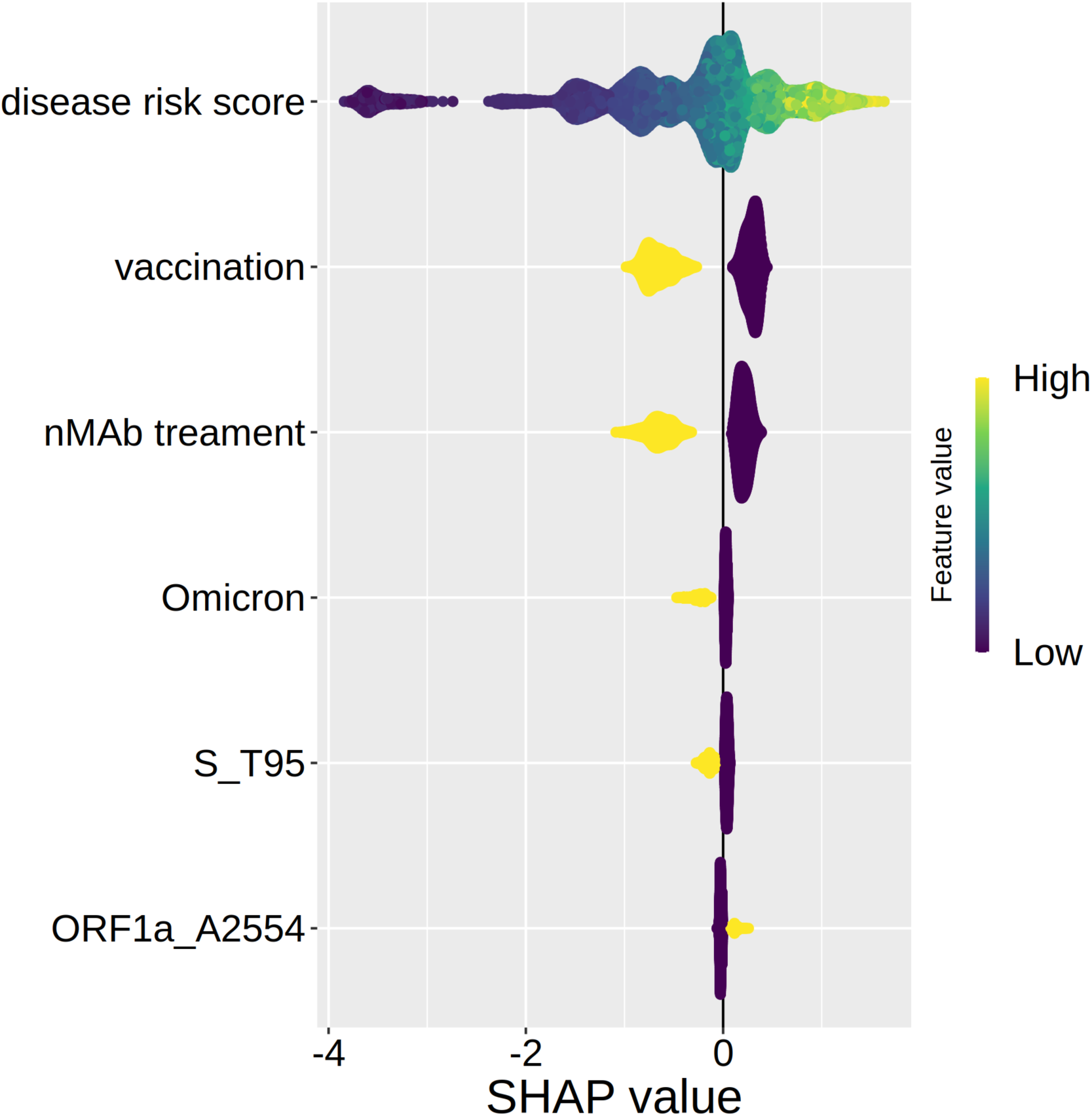
Distribution of SHAP values for the six features with highest mean absolute SHAP values. Each point represents the median SHAP value for one sample across the 25 data split-folds. Colors indicate the magnitude of the feature value for each sample. All features, apart from disease risk score, are Boolean. SHAP values are in log odds ratio units and indicate the contribution a feature makes to the predicted hospitalization risk for each sample compared to the mean hospitalization rate.

**Table 1.**
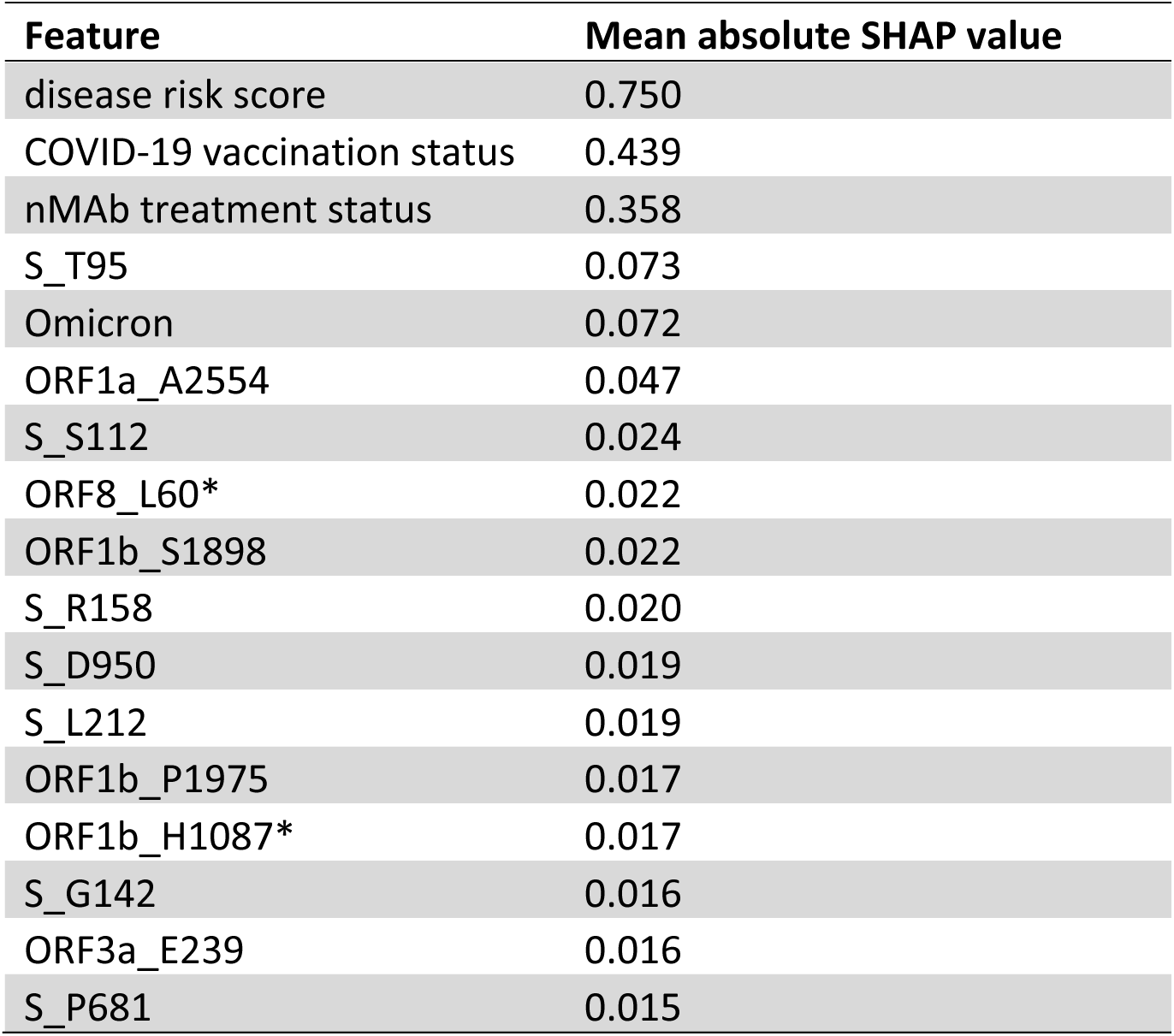
Features with mean absolute SHAP values above 0.015, calculated from 25 data split-fold combinations. *ORF8_L60 and ORF1b_H1087 alternate alleles are correlated with each other and with ORF1a_H2125 at greater than 0.9 in this dataset. All other genetic variants in this table have no correlations with other genetic variant features greater than 0.9.

| Feature | Mean absolute SHAP value |
| --- | --- |
| disease risk score | 0.750 |
| COVID-19 vaccination status | 0.439 |
| nMAb treatment status | 0.358 |
| S_T95 | 0.073 |
| Omicron | 0.072 |
| ORF1a_A2554 | 0.047 |
| S_S112 | 0.024 |
| ORF8_L60* | 0.022 |
| ORF1b_S1898 | 0.022 |
| S_R158 | 0.020 |
| S_D950 | 0.019 |
| S_L212 | 0.019 |
| ORF1b_P1975 | 0.017 |
| ORF1b_H1087* | 0.017 |
| S_G142 | 0.016 |
| ORF3a_E239 | 0.016 |
| S_P681 | 0.015 |

The genetic features with the largest impacts on hospitalization predictions were an amino acid change in the spike (S) protein at position 95 (S_T95), the Omicron WHO variant, and an amino acid change in open reading frame 1a at position 2554 (ORF1a_A2554) (see Table 1 and Figure 2). Both an alternate allele at S_T95 and Omicron status, i.e., infection with an Omicron strain versus a strain of any other WHO variant, predicted lower probability of hospitalization. There was just one substitution at S_T95 in this dataset, isoleucine (I) replacing threonine (T); this occurred in 3,595 samples, including in nearly all Omicron and Mu samples as well as in 14% of the Delta samples. An amino acid change at ORF1a_A2554, which occurs in non-structural protein 3 (nsp3) at amino acid position 1736, predicted slightly increased hospitalization risk. All but one of the samples with an amino acid change at this site had valine (V) substituted for alanine (A). Amino acid substitutions at this site occurred in 1,902 samples, including 23% of Delta samples and 3% of Omicron samples. Of the 14 genetic variant features shown in Table 1, seven are in the spike protein, and five of these are in the spike protein’s N-terminal domain.

A SHAP analysis of feature interactions indicates that the effects of several genetic variables on predicted hospitalization risk varied depending on patient covariate values. Table 2 shows the pairs of features with the highest mean SHAP interaction values. Omicron status had mean SHAP interaction values exceeding 0.02 with disease risk score, COVID-19 vaccination status, and nMAb treatment status. Figure 3 shows the impact of Omicron status on hospitalization predictions by disease risk score, COVID-19 vaccination status, and nMAb treatment status. SHAP values are lower than zero for patients infected with Omicron strains, indicating that being infected with Omicron instead of another WHO variant predicted reduced odds of hospitalization. In addition, the predicted impact of Omicron status in reducing hospitalization was greatest in unvaccinated, non-nMAb treated patients, while Omicron status had less impact on predicted hospitalization in those known to be vaccinated and nMAB-treated. Furthermore, the predicted impact of Omicron status was greatest for those with intermediate disease risk score values. SHAP values for S_T95 and S_S112 also varied depending on patient covariates, and their relationships with disease risk score, COVID-19 vaccination status, and nMAb treatment status are shown in Figures S1-S2 in Supplemental Materials. Finally, COVID-19 vaccination and nMAb treatment both had substantial interactions with disease risk score, and COVID-19 vaccination and nMAb treatment interacted with each other. The effect nMAb treatment on predicted hospitalization tended to increase with disease risk score, while the effect of COVID-19 vaccination on predicted hospitalization was greatest in those with intermediate disease risk score values (Figures S3-S4 in Supplemental Materials).

**Figure 3.**
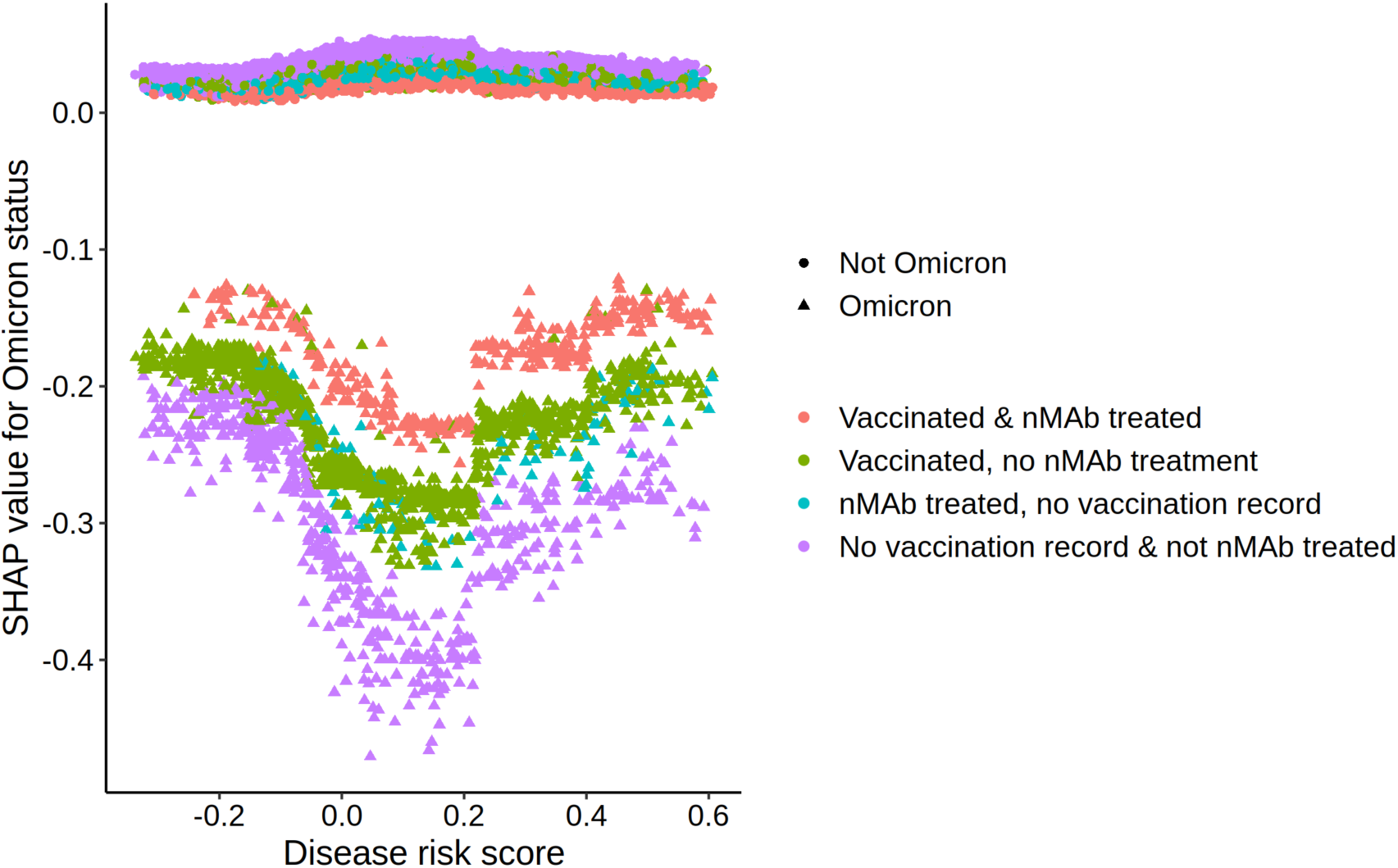
Median SHAP values for Omicron status for all samples by disease risk score, COVID-19 vaccination status, and nMAb treatment status. Omicron samples are represented by triangles and non-Omicron samples are represented by circles. SHAP values above zero indicate an increase in predicted risk of hospitalization, while those below zero indicated a decrease in predicted risk of hospitalization.

**Table 2.**
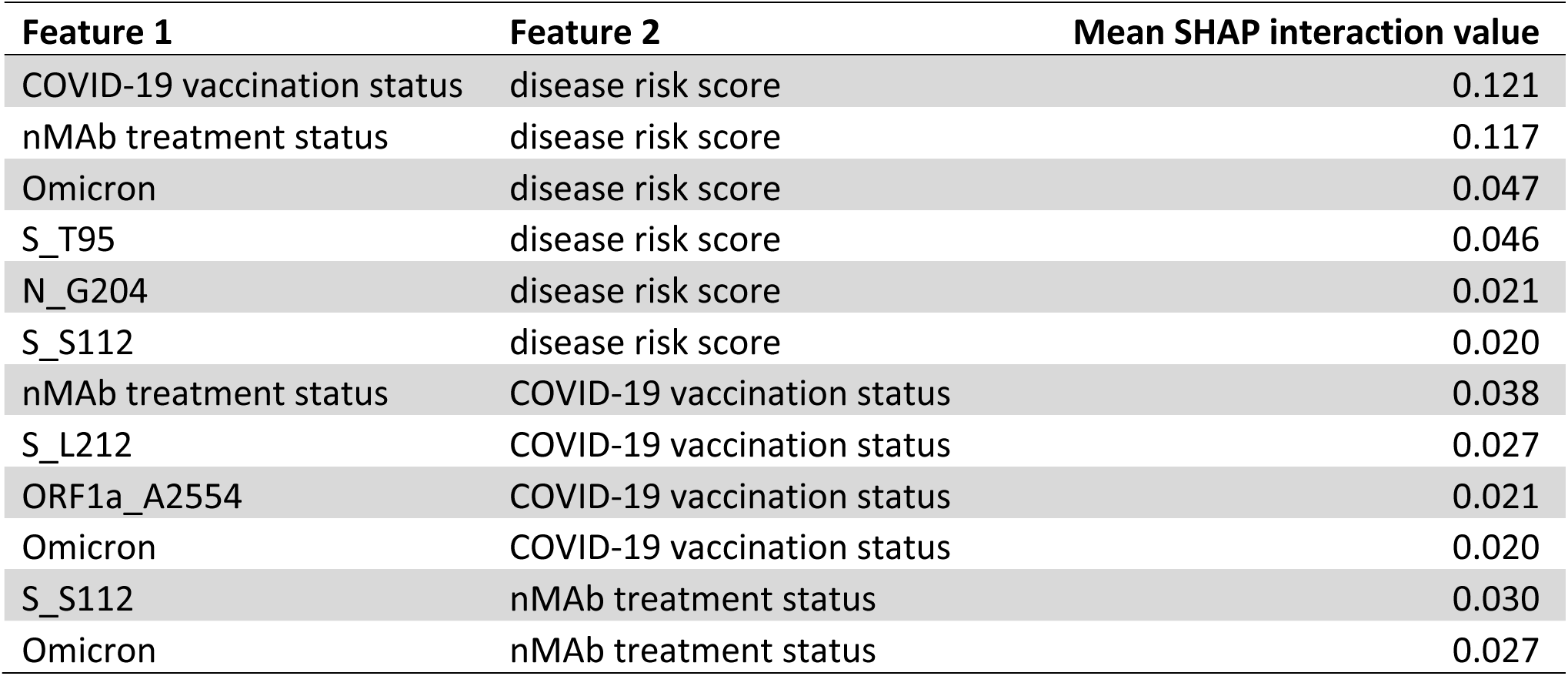
Median SHAP interaction values from 25 split-folds for pairs of variables with the interaction value exceeding 0.02.

### GLMM modeling results

GLMM were fitted using stochastic variational inference for each of the 28 amino acid changes at the sublineage level. In each case, model fitting resulted in posterior distributions for all model parameters. The results presented below are similar to those for other choices of priors (see Supplementary Materials Table S4). Figures S5 and S6 in Supplementary Materials demonstrate that the GLMM model correctly approximates risk for the hospitalization outcome.

Representative posterior distributions for the *α*_1_, *α*_2_, and *α*_3_ parameters (corresponding to nMAb, disease risk score, and COVID-19 vaccination status effects, respectively) are shown in Figure 4. The posterior distributions are shown on the logit scale. As expected, treatment with nMAb or having been at least partially vaccinated is associated with lower odds of hospitalization, with posterior distributions of these parameters lying entirely below zero. Also, a higher disease risk score is associated with higher odds of hospitalization, as demonstrated by that posterior distribution lying entirely above zero.

**Figure 4.**
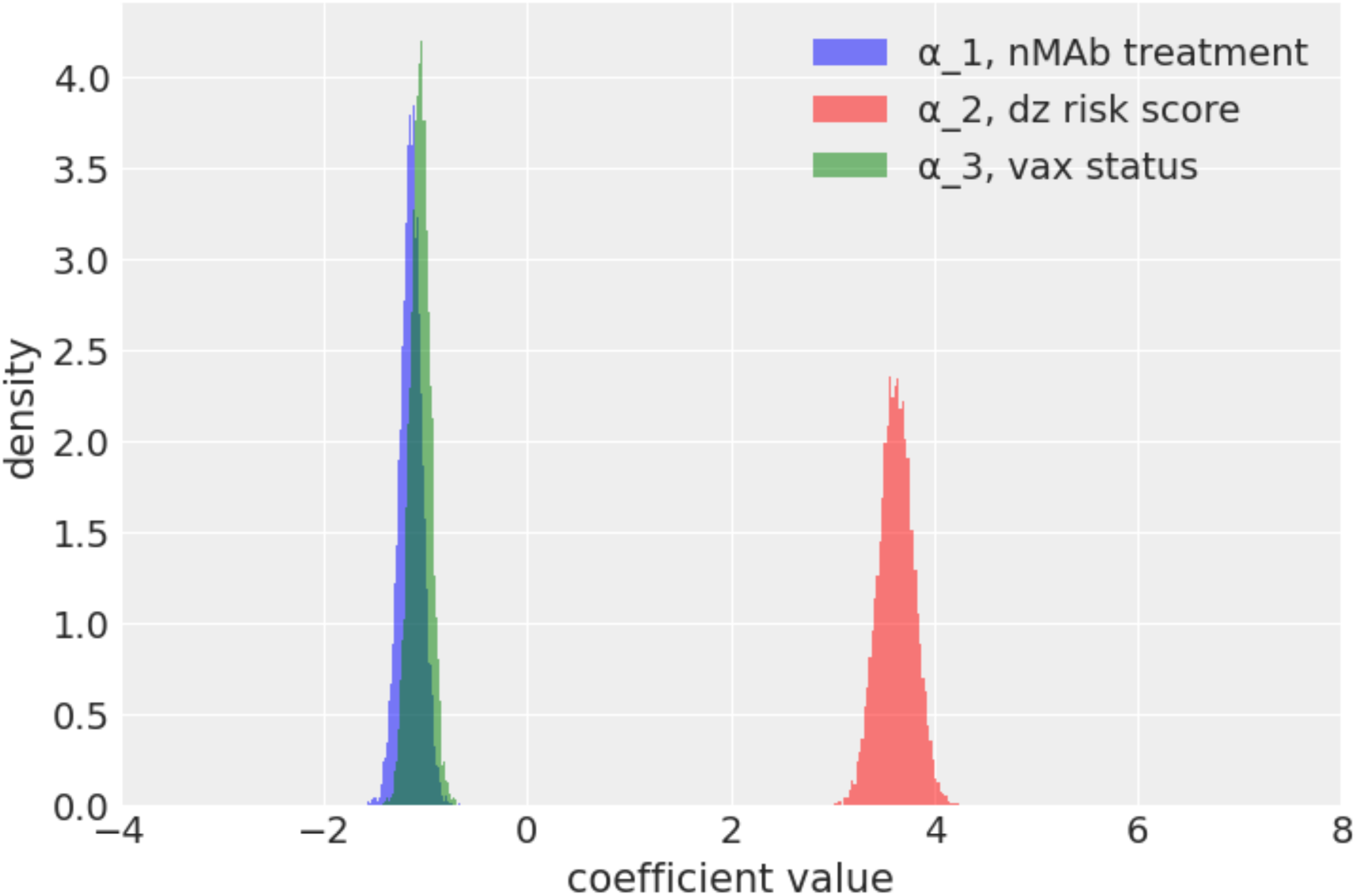
Representative posterior distributions from Bayesian mixed modeling for the *α*1, *α*2, and *α*3 parameters, corresponding to nMAb, disease risk score, and COVID-19 vaccination status effects, respectively. The posterior distributions are shown on the logit scale. Results are shown for a model with one specific amino acid change and are representative of results for models with other amino acid changes.

Figure 5 shows representative posterior distributions for the lineage-specific intercept estimates for the 30 most common lineages (each represented by at least 50 samples). Each line in this forest plot represents the posterior distribution of the lineage intercept estimate of the correspondingly labeled lineage. The baseline expectation, corresponding to no lineage effect (with a value equal to the overall risk of admission on the logit scale) is shown as a vertical dashed red line. The width of the posterior reflects residual uncertainty after model fitting. Mean values of each posterior are shown as an open circle on each line. The figure suggests a range of associations between lineage and hospitalization risk, with lineages such as BA.1 being associated with lower risk, AY.26 with average (i.e., baseline) risk, and B.1.621 with higher hospitalization risk. Of note, several of the lineages seemingly associated with lower risk are Omicron lineages (e.g., BA.1, BA.1.20, BA.1.18, BA.1.1); this is consistent with the predicted effects of the Omicron feature seen in the XGBoost/SHAP analysis (see Table 1 and Figure 2 above).

**Figure 5.**
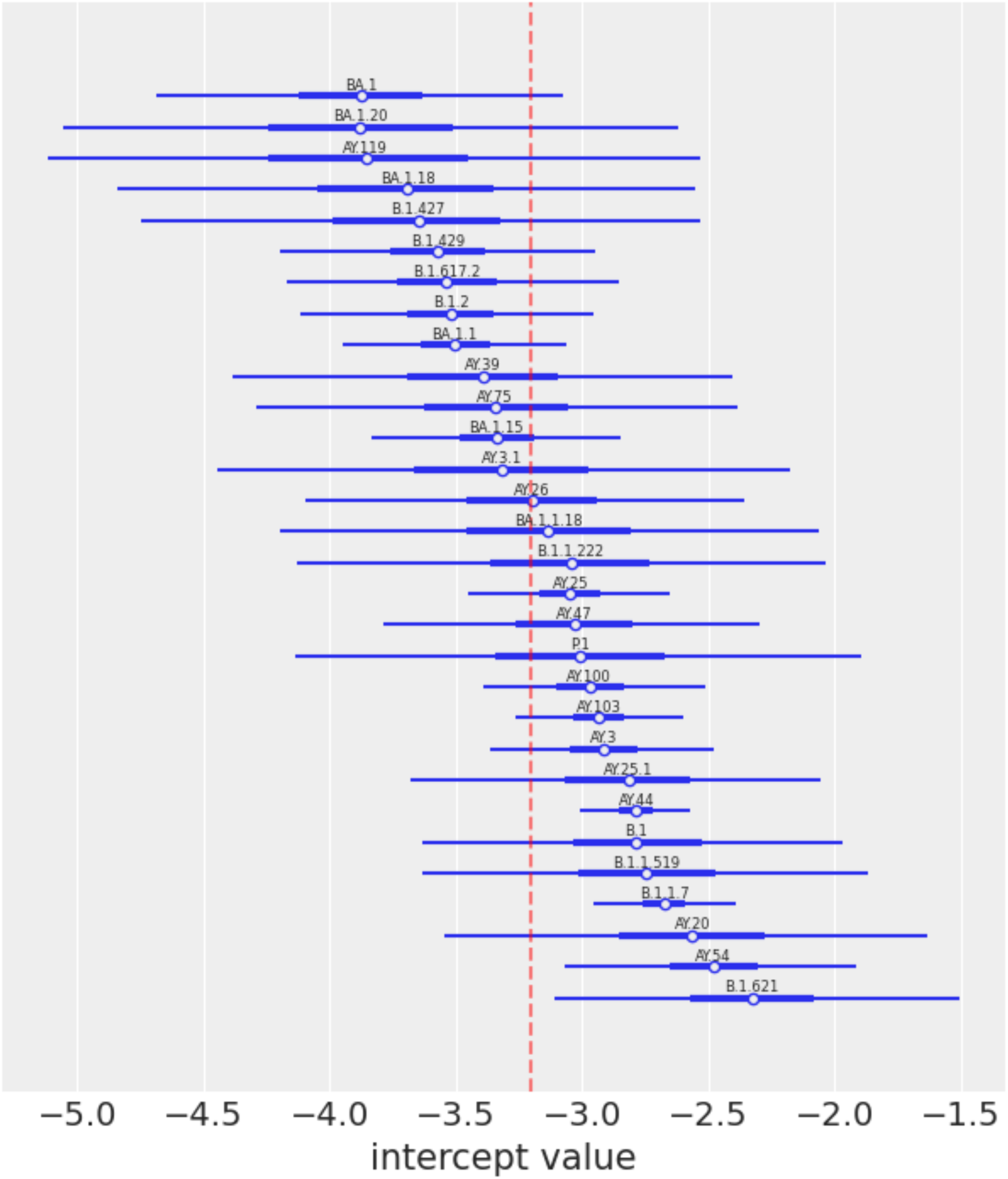
Representative posterior distributions for the lineage-specific intercept estimates for the 30 lineages with at least 50 samples. Each line in the forest plot represents the posterior distribution (the highest posterior density interval for 94% of density) of the lineage intercept estimate for each correspondingly labeled lineage. The interquartile range and mean of each posterior correspond to the thicker part of each line segment and the open circle, respectively. The baseline expectation, corresponding to overall risk of admission on the logit scale, is shown as a vertical dashed red line.

Table 3 shows quantitative evidence that as many as six of these lineage intercept estimates may be different from the baseline expectation, i.e., no lineage effect. These lineages were identified by posterior distributions with extreme z-scores (> 1.6 or < -1.6) or with Bayes factors > 3. In the case of AY.44, the Bayes factor is very large, with a value greater than two million. This indicates that the posterior distribution is very different from the prior, which can also be seen in Figure 5.

**Table 3.** Lineages potentially associated with hospitalization according to GLMMs (either |z-score| > 1.6 or Bayes factor > 3).

| Lineage | Lineage frequency | Number of samples | Z-score | Bayes factor |
| --- | --- | --- | --- | --- |
| AY.103 | 0.108 | 1352 | 1.80 | 0.70 |
| AY.44 | 0.153 | 1922 | 4.18 | 2006632.20 |
| AY.54 | 0.014 | 172 | 2.81 | 10.01 |
| B.1.1.7 | 0.087 | 1090 | 4.16 | 213.07 |
| B.1.621 | 0.006 | 69 | 2.44 | 7.13 |
| BA.1 | 0.021 | 268 | -1.86 | 2.00 |

Table 4 shows amino acid changes highlighted by Bayes factors > 3 (none were identified with extreme z-scores). These amino acid changes have at least some evidence of association with hospitalization risk according to the GLMMs. The analyses indicate that alternate amino acids at ORF1b_P1975, ORF1a_R4179, S_L212, and S_S112 may be associated with increased risk of hospitalization (positive z-scores), while alternate amino acids at the other seven amino acid change sites may be associated with decreased risk for hospitalization (negative z-scores). Note that for amino acid change S_G142, two associations are highlighted, one within lineage AY.3, and a second within AY.100; in both cases, the amino acid change appears to be negatively associated with hospitalization risk. Results with different priors consistently identified the same set of amino acid changes (see Supplementary Materials Table S4). Also of note, most of the amino acid changes highlighted by GLMMs were also in the top ranked features identified in the XGBoost-SHAP analysis; specifically ORF1b_P1975, S_D950, S_G142 S_L212, and S_S112 were found in both approaches (Table 1).

**Table 4.**
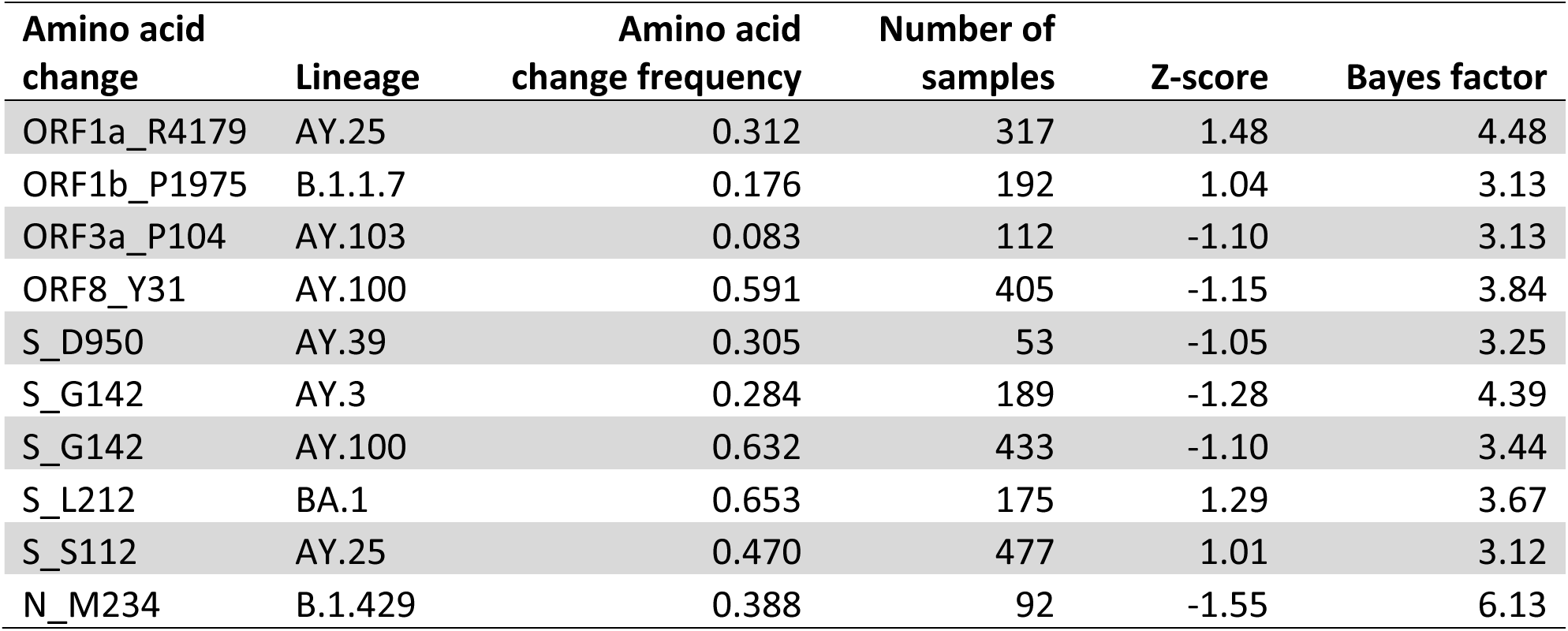
Amino acid changes potentially associated with hospitalization according to GLMMs. None were identified as having extreme z-scores (|z-score| > 1.6), instead all were identified because their Bayes factors were > 3. Similar results were found for a range of priors on *σ*_1_ (see Supplementary Materials Table S4).

| Amino acid change | Lineage | Amino acid change frequency | Number of samples | Z-score | Bayes factor |
| --- | --- | --- | --- | --- | --- |
| ORF1a_R4179 | AY.25 | 0.312 | 317 | 1.48 | 4.48 |
| ORF1b_P1975 | B.1.1.7 | 0.176 | 192 | 1.04 | 3.13 |
| ORF3a_P104 | AY.103 | 0.083 | 112 | -1.10 | 3.13 |
| ORF8_Y31 | AY.100 | 0.591 | 405 | -1.15 | 3.84 |
| S_D950 | AY.39 | 0.305 | 53 | -1.05 | 3.25 |
| S_G142 | AY.3 | 0.284 | 189 | -1.28 | 4.39 |
| S_G142 | AY.100 | 0.632 | 433 | -1.10 | 3.44 |
| S_L212 | BA.1 | 0.653 | 175 | 1.29 | 3.67 |
| S_S112 | AY.25 | 0.470 | 477 | 1.01 | 3.12 |
| N_M234 | B.1.429 | 0.388 | 92 | -1.55 | 6.13 |

## DISCUSSION

In this work, clinical and SARS-CoV-2 viral genomic data from 12,538 patients was examined for association of viral genomic variants with hospitalization within 14 days of COVID-19 diagnosis. We created and used a model-based disease risk score, summarizing risk from dozens of co-morbidities and demographic factors, and incorporated information on COVID-19 vaccination and nMAb treatment status, thus accounting for their influences on outcome (23). Using two different modeling approaches, we find evidence for the association of several amino acid variants with hospitalization. To our knowledge, this study is the largest to date that combines detailed clinical and viral genomic information with the clinical outcome of hospitalization, and the only such study that uses data from multiple health systems.

The results of this study are consistent with and extend the results of the prior study from which this study’s dataset is derived (23). Like in the prior study which used a different analytical method, both the XGBoost-SHAP and GLMM analyses identified COVID-19 vaccination and nMAb treatment as predictive of lower risk of hospitalization. This study found that SARS-Co-V-2 genetic factors were also predictive of hospitalization in this dataset. While the genetic factors, including WHO variant, collectively improved prediction in the XGBoost models, the individual contributions of WHO variants and amino acid mutations were small compared to the magnitude of the effects of the patient covariates. Furthermore, the SHAP interaction results suggest that the associations of these genetic variants with hospitalization risk may vary depending on patient condition.

This study used two different modeling approaches to investigate genetic associations at different scales. GLMMs were used to investigate variation among lineages and amino acid variation within lineages, while XGBoost with SHAP analysis was used to investigate amino acid variants across lineages and interactions between genetic variants and patient covariates. Because these analyses investigated associations in different subsets of the data, the finding of overlapping but not identical sets of top candidate variants is not surprising. Nevertheless, five of the nine candidates from the GLMM were observed in the top 14 genetic features in the XGBoost-SHAP analysis. S_T95, the top XGBoost-SHAP genetic feature, was not evaluated in the GLMMs because it did not meet the frequency and number thresholds in any one lineage but was detected in the XGBoost-SHAP analyses due to its occurrence in multiple lineages representing three different WHO variants. Similarly, the following amino acid changes highlighted by the XGBoost analysis were not included in GLMMs because they did not meet frequency and number thresholds in any one lineage: ORF1a_A2554, ORF1b_S1898, ORF3a_E239, and S_P681.

Of the 14 genetic features with mean absolute SHAP values above 0.15, half occurred in the spike protein. Five of these, S_T95, S_S112, S_G142, S_R158, and S_L212, occur in the N-terminal domain (NTD), S_P681 occurs in the S protein cleavage site, and S_D950 occurs in the C-terminal domain. Interestingly, none of the genetic features were found in the S protein receptor binding domain (RBD). A few of these have been associated with disease severity in prior studies. Sokhansanj et al. identified mutations at S_G142 as contributing to prediction of disease severity in a deep learning model using S protein features (8), and Liang et al identified both S_T95 and S_L212 as associated with COVID-19 disease severity in a study of associations between spike amino acid mutation prevalence in GISAID data and concurrent hospitalization rates in CDC data (7). Obermeyer et al. also found that the amino acid substitution S T95I was associated with increased SARS-CoV-2 transmission (22), and other studies point to associations of S_T95 with increased transmission (47). Shen et al. reported that S_T95I was associated with increased viral loads in patients, based on PCR cycle threshold results, and that S_T95I arose multiple times in Delta lineages in addition to its presence in Omicron strains (48). Together, these results suggest that the S_T95I mutation may increase transmission while reducing disease severity. Within the spike protein (S), the polar threonine amino acid at position 95 (S_T95) resides within a five-angstrom pocket of hydrophobic residues (S_F186, S_L189, S_I210, S_L212, S_A264). The change from the polar tyrosine residue to the hydrophobic isoleucine residue (S_T95I) could conceivably result in a stabilization of the globular region that S_T95I resides within (see Figure 8).

**Figure 8.**
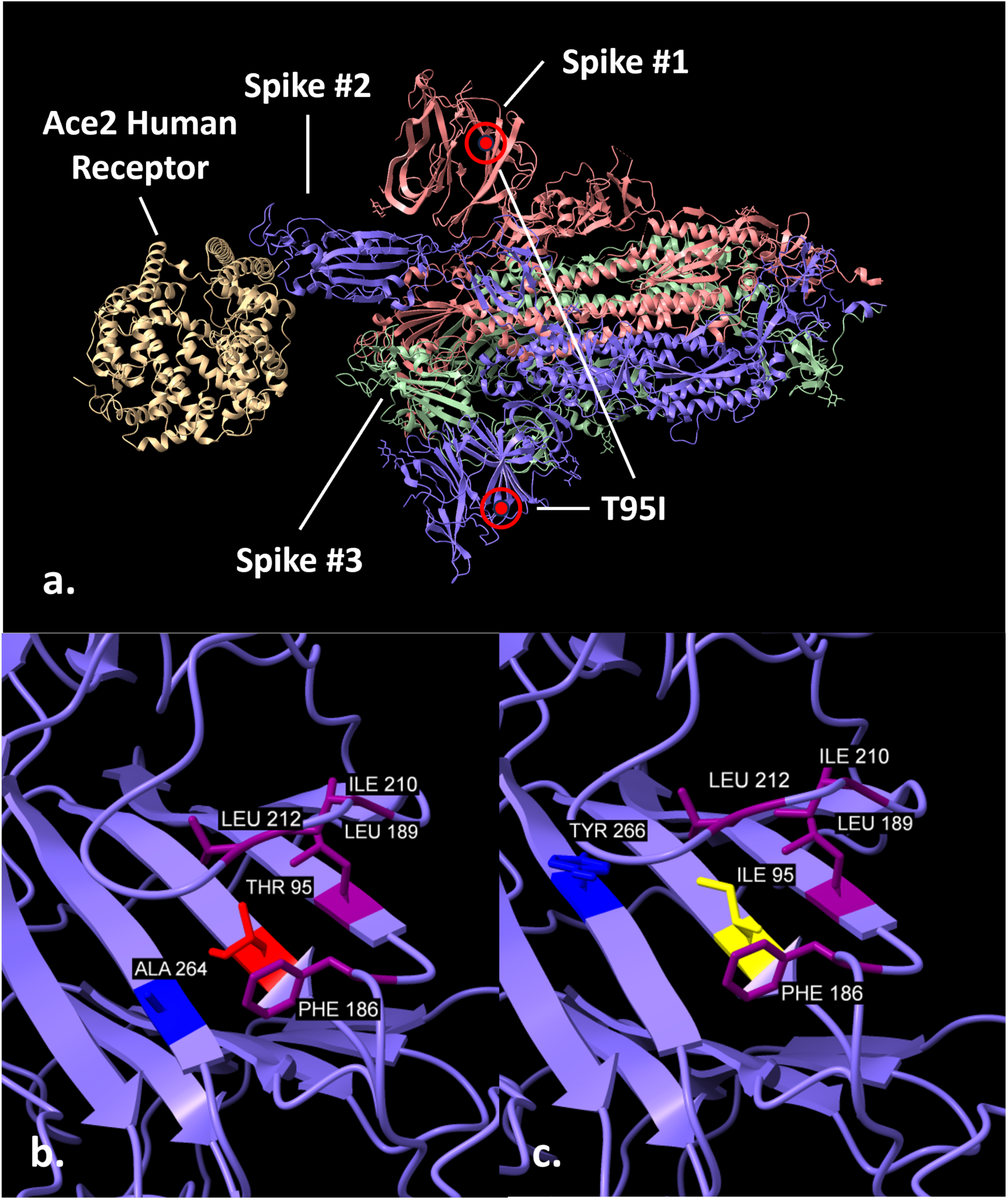
Location and environment of the T95I mutation. The hydrophobic ILE 95 substitutes for the polar THR 95 in the hydrophobic pocket of surrounding residues. All residues shown are within five angstroms of the 95 residue atoms.

The GLMM analysis identified several candidates, including one in the Alpha lineage, ORF1b_P1975 (in nsp14 at position 451). The most common genetic mutation for ORF1b_P1975, C19390T, was also found to be weakly associated with severe COVID-19 in a prior GWAS study (14). While we are not aware of papers about this mutation’s functional effects, nsp14 has been found to shut down host protein synthesis and thereby interfere with host innate immune responses (49).

Other candidate variants found by two or more prior studies include nsp6 amino acid variant L37F, which was associated with asymptotic or mild symptoms (2, 6, 9); spike protein (S) variant V1176F (2, 4) and open reading frame 3a (ORF3a) variant Q57H (2, 10), which were associated with more severe symptoms; and S variant D614G, which was associated with more severe symptoms in three papers (2, 10, 11) and less severe outcomes in another paper (3). None of these were identified in our study. ORF1a_L37F and ORF3a_Q57H were tested in our dataset, and S_V1176F was part of the set of amino acid changes highly correlated with Gamma; however, S_D614 was not tested because it fell outside the minimum allele frequency threshold in our dataset. One possible reason for differences in associations is that our study used a different measure of disease severity and a different study population–hospitalization in persons with co-morbidities. In contrast, many of the prior studies have used symptom information for severity (6, 16), evaluated differences in symptoms among patients that were already hospitalized (13, 14), or included persons without co-morbidities (3, 7).

This study has several important limitations. First, the genetic variants we highlight represent correlations, not causation, with our outcome. As discussed above, there are biologically plausible reasons that could explain some of the associations, however we cannot eliminate the possibility that they represent false positive findings. The fact that some of the variants we highlight have been observed in previous studies supports the notion that they are not spurious, but further research would be needed to claim causality.

Second, we used all-cause hospitalization within 14 days of diagnosis of COVID-19 as our outcome of interest. Other possible outcomes we could have explored include death within 14 or 30 days of diagnosis, or hospitalization within 30 days of diagnosis. Hospitalization within 14 days is a rare outcome in our dataset, and the limited number of cases hinders our ability to detect associated genomic variants. Death, either within 14 or 30 days, is even rarer (0.1-0.3% of patients), and thus, we did not use this outcome. By limiting to 14 days, our aim was to enrich for hospitalizations that are truly related to severe COVID-19 illness. However, it is likely that a small fraction of these hospitalizations was not COVID-19 related, and thus represent misclassifications. This problem would likely be worse had we used 30-day hospitalization.

Third, although this dataset is relatively large, statistical power is a limitation. This is due to the low rate of hospitalizations discussed above, the complexity of the viral genomic data (e.g., many different lineages) and the clinical data, and the relatively small magnitude of the associations observed between viral genomic variants. A larger sample size would likely help clarify which findings represent false positives and false negatives.

Fourth, the data used here were collected primarily to support an observational cohort study of the effectiveness of nMAbs for COVID-19 (23), and genomic data from some samples did not have a high read depth. As such, aspects of the viral genomic data were limiting. For example, viral genomic data from 1,259 samples were discarded as part of quality-screening procedures in the original cohort study (24). Data from an additional 1,165 samples were discarded as part of the Nextclade quality threshold set for the present study. Even after applying these quality thresholds, various sequence gaps and ambiguities remained. To address these, we imputed missing / ambiguous base calls by placing our samples on a large, publicly available SARS-CoV-2 phylogenetic tree using UShER. While most placements on the tree were unambiguous, some were not, and misplacements are likely to have resulted in some incorrect imputed values in the dataset used for analyses.

Fifth, this study investigated only the presence of amino acid substitutions or stop codons at sites. Synonymous nucleotide variation that has effects beyond protein component could be important, as could variation caused by deletions. Our approach pooled both different nucleotide substitutions that cause the same amino acid substitution, and furthermore all alternate amino acid substitutions at a site. This was done to increase power to detect associations by increasing the number of samples with a given feature, and to jointly analyze amino acid changes that occurred in different lineages and WHO variants via different nucleotide or amino acid substitutions. This pooling approach may obscure varying associations for different amino acid substitutions.

Finally, while we evaluated the overall contribution of genetic variation in both the XGBoost and GLMM models, we have not evaluated the statistical significance of individual amino acid changes. For the XGBoost-SHAP analysis, methods for evaluating statistical significance are an active area of research (50). In the case of GLMMs, we ran separate versions of the model for each of 28 amino acid changes and did not apply multiple-testing correction methods. It is possible that some of the associations we have highlighted are due to chance. However, the number of tests is relatively small number compared to most GWAS. Moreover, these 28 tests are not truly independent, since there exist varying degrees of correlation within this set of 28 amino acid changes; for example, three amino acid changes in this set, ORF1b_H1087, ORF1a_H2125, and ORF8_L60, each have pair-wise Pearson correlations greater than 0.9.

The COVID-19 pandemic has highlighted the need for vigilance in the face of possible future pandemics. Large-scale linked clinical-genomic datasets are one possible approach to uncovering associations that may lead to improved pathophysiologic understanding and, perhaps, therapeutic targets. As such, enduring capabilities that combine clinical and genomic data from multiple sites within a secure infrastructure, like the dataset used in this study, should be considered as possible measures for ongoing national pandemic preparedness.

## Data Availability

Deidentified data has been submitted to National COVID Cohort Collaborative (N3C). Available at https://ncats.nih.gov/n3c.

## ACKNOWLEDGEMENTS

We thank Fraser W. Gaspar and Steven Z. Fairchild of the MITRE Corporation for helpful feedback on this manuscript.

The authors wish to acknowledge the following persons, who each played an important role in the parent observational cohort study of the effectiveness of neutralizing monoclonal antibodies for COVID-19. From the MITRE Corporation: Nalini Ambrose; Brian Anderson; Francis Campion; Risa Danan; Lauren D’Arinzo; Heath Farris; Kristin Fitzgerald; Melissa Garcia; Fraser W. Gaspar; Eric Neumann; Nathan Welch; Jennifer Yttri. From the University of California, Irvine: Alpesh Amin; Daniel Chow; Suzanne Sandmeyer. From the Biomedical Advanced Research and Development Authority (BARDA): Julio Barrera-Oro; Karl Erlandson. From Harvard Medical School: Monica Bertagnolli; Steven Piantadosi. From Houston Methodist: Ashley Drews; Stephen Jones; James Musser; Farhaan Vahidy. From Booz Allen Hamilton in support of BARDA: Carlene Gong. From Tunnell Government Services in support of BARDA: George Hanna. From Intermountain Health: Bert Lopansri; Brandon Webb. From Mayo Clinic: John O’Horo; Bobbi Pritt; Raymund R. Razonable.

## Competing Interests

There are no competing interests to declare.

## Funding

The work presented in this study was supported solely by MITRE’s Independent Research and Development Program.

## Notes

### Competing Interest Statement

The authors have declared no competing interest.

### Author Declarations

The institutional review board at MITRE determined that the study qualified for exempt status under the provisions of 45 CFR Part 46.104(d)(4).

