## Supplementary Materials for "SARS-CoV-2 Genetic Variants and Patient Factors Associated with Hospitalization Risk"

**Supplementary Text**

**nMAbs Dataset**

Details of the nMAbs dataset have been described elsewhere (23). Briefly, the nMAbs study involved collecting structured, deidentified patient-level data from four health systems within the United States: Houston Methodist, Intermountain Health, Mayo Clinic, and University of California, Irvine. Data were shared to a secure, centralized repository for cleaning and analysis. Each patient was associated with an index date, representing the day of first known laboratory-confirmed COVID-19 diagnosis. This index date was designated "day 0"; the time of all patient clinical events was then expressed as days relative to the index date. Although actual calendar dates were not available, the month and year of the index date were known for each patient. The institutional review boards at MITRE and the four health systems determined that the study qualified for exempt status under the provisions of 45 CFR Part 46.104(d)(4).

Structured clinical data shared as part of the nMAbs study included patient demographic information, diagnosis / condition information (most commonly ICD10-CM codes), procedures (ICD10-PCS, CPT, or local codes), measurements (e.g., lab tests / results, associated with LOINC codes), encounters (e.g., hospitalizations, ambulatory clinic visits, emergency department visits), observations (e.g., vital signs, patient-level assertions), drug exposures (including inpatient and outpatient pharmacologic treatments, often represented as RXNORM codes; and including vaccinations, represented by CVX codes). Inclusion criteria for the nMAbs study were as follows: patients aged 12 years or older, laboratory-confirmed COVID-19 diagnosis (via nucleic acid amplification or antigen test, or if the patient was referred specifically for nMAbs treatment), diagnosed in a non-inpatient setting from November 2020 to January 2022, and at least one FDA nMAbs Emergency Use Authorization "high risk" criterion. Patients were excluded if they died on the index date, had evidence of a prior COVID-19 infection, received an outpatient COVID-19 treatment other than nMAbs, or if they were treated with nMAbs > 10 days after diagnosis. The final nMAbs cohort had 167,183 patients; 13,703 of these patients had linked viral genomics data (see below). The outcome explored in this study was all-cause hospitalization within 14 days of day 0; any person with an inpatient encounter between day 1 and day 14 (inclusive) was counted as "hospitalized". In the full set of 167,183 patients, the overall fraction of patients hospitalized within 14 days of COVID-19 diagnosis is 0.031, the fraction of patients with any record of COVID-19 vaccination prior to COVID-19 diagnosis is 0.372, and the fraction of patients treated with nMAbs is 0.151. Further details on nMAbs data, including data missingness and imputation strategies, can be found in (23).

**Genomics Data Pipeline**

Details of the nMAbs genomics pipeline have been described elsewhere (24). Briefly, viral genomics data derived from 16,055 biologic specimens were shared to the centralized repository for the nMAbs study. After processing and filtering for data quality and other issues (see below), data from 14,796 samples remained. After linking samples to patient structured clinical data discussed above, data from 13,703 quality filtered samples were

© 2024 The MITRE Corporation, All Rights Reserved
Approved for Public Release. Distribution Unlimited. Public Release Case Number 24-0748.

available from patients meeting inclusion criteria for the nMAbs study. We retained viral genomic data for only one sample per patient, taken on each patient's index date.

Viral genomics data were submitted as FASTQ files. In all, 24,887 FASTQ files (7,223 files from single-end read runs and 17,664 paired files from paired-end read runs) were submitted. The genomics pipeline processed these files in a series of discrete steps. After ensuring that human reads and any potential personally identifiable information were excluded, paired FASTQ files were analyzed together, and single FASTQ files individually. FASTQ files were quality trimmed and aligned to the reference SARS-CoV-2 genome (NCBI Reference Sequence NC_045512.2). Pileup files were created and used as inputs for variant caling and annotation (via iVar variants (51) and SnpEff (52), respectively). Pileup files were also used to produce consensus FASTA files.

A set of quality criteria were devised to ensure that only data from high-quality samples were retained for analysis. These criteria included: 1) consensus FASTA length >= 27,000 bases; 2) number of ambiguous base calls <= 5,000; 3) rate of base calls with a quality Phred score of at least 30 >= 80%; and 4) percentage of genome covered at least 10X >= 90% (or >= 80% if criteria 1, 2, and 3 were all satisfied). Additional steps were taken to identify missing base calls due to gaps as described in (24), and the criteria for retaining base calls in the dataset were: 1) minimum allele frequency of 0.75, 2) minimum quality threshold of 30, and 3) minimum depth of 10.

Within the set of samples passing all data quality checks, there were still gaps in genomic sequence coverage and ambiguous base calls (24). To address these, we used UShER (30) to place all of our samples on the existing public phylogenetic tree of SARS-CoV-2 genomes with maximum parsimony. By considering the entire consensus genome sequence for each sample, UShER placement can essentially suggest likely possibilities for missing or ambiguous base calls.

The consensus fasta genome sequences for all samples were compiled into one multifasta file and used as input to UShER, along with the public tree from December 2022 (53). We used the default setting of 1 for the –multiple-placements parameter. Each of our samples was thus associated with a node on the public tree, or to a new node with unique nucleotide substitutions just under a public tree node. Reading off these nodes gives an expected set of single nucleotide substitutions for each sample (based on the `mutation-paths.txt` UShER output file). Although each sample was only placed on one node in the tree, UShER reports a count of equally optimal placements for each sample. For our original consensus fasta sequences, 57% of samples had exactly one optimal placement, 81% of samples had 1, 2, or 3 equally optimal placements, and 93% of samples had <= 10 optimal placements. When there were multiple optimal placements, the node chosen by UShER was used, i.e., the node with the greatest number of descendant leaves (30).

The original consensus fasta genome sequences were also processed by the Nextclade CLI 2.12.0 tool (32) to obtain insertion / deletion calls. We accepted indel calls from Nextclade if sequence quality was flagged as "good" or "mediocre" if insertions did not contain ambiguous bases. For samples with "bad" quality, we assumed the presence of indels characteristic for the lineage the sample was assigned to by Nextclade. We identified indels

© 2024 The MITRE Corporation, All Rights Reserved
Approved for Public Release. Distribution Unlimited. Public Release Case Number 24-0748.

characteristic of each Nextstrain clade that appeared in the dataset using cov-spectrum (54) and corroborated that these indels were consistent with the indels in the "good" samples. One clade, 21A, had diversity within that segregated by Pango lineage; consequently, reference indels were set for groups of Pango lineages within this clade. We also performed some indel clean up that included replacing ambiguous nucleotides in common insertions and adjusting deletion calls that were one position off for common deletions.

Based on the final position within the SARS-CoV-2 phylogenetic tree and the deletion mutations, we created new consensus fastas for all samples by incorporating all these mutations as changes relative to the reference Wuhan genome (55). We refer to these updated consensus sequences as UShER-cleaned sequences. We applied Nextclade to these sequences to compile a set of expected mutations for each sample.

To mitigate the possibility of incorrect imputation of viral genomic sequence using UShER, we filtered based on the quality of the UShER-cleaned sequences, using the Nextclade "overall quality score". We established a threshold of 250 for this score, thus retaining 12,538 samples with higher quality for further analysis (91.5% of the full set of 13,703 samples). The most common quality issue flagged by Nextclade within the set of 12,538 samples retained for analysis was "unlabeled private mutations".

**Amino Acid Variants in the Analyses**

The following amino acid changes were included as features in the XGBoost-SHAP analyses. The 28 marked with asterisks (*) were also analyzed using GLMMs: N_A208*, N_D343, N_G18, N_G204, N_G215, N_M234*, N_P199, N_P67, N_R203, N_T205, ORF1a_A1306, ORF1a_A2554, ORF1a_A3209, ORF1a_A599*, ORF1a_C3766, ORF1a_D1228, ORF1a_E1633, ORF1a_G519, ORF1a_H2125*, ORF1a_I3731, ORF1a_I4205, ORF1a_K3353, ORF1a_L3352, ORF1a_L3606, ORF1a_M2259*, ORF1a_M2606*, ORF1a_P1640*, ORF1a_P2046*, ORF1a_P2287, ORF1a_P309, ORF1a_P959, ORF1a_Q3966*, ORF1a_R4179*, ORF1a_T265, ORF1a_T3255, ORF1a_T3646, ORF1a_T3750, ORF1a_V2930, ORF1a_V3718, ORF1b_A1219, ORF1b_A1918, ORF1b_H1087*, ORF1b_I1257, ORF1b_K2557*, ORF1b_N1653, ORF1b_P1570, ORF1b_P1975*, ORF1b_Q2635, ORF1b_R2613, ORF1b_S1898, ORF3a_E239, ORF3a_G172, ORF3a_L106, ORF3a_P104*, ORF3a_Q57, ORF6_K48, ORF7a_V71, ORF7b_T40, ORF8_C25*, ORF8_C61*, ORF8_L60*, ORF8_P36*, ORF8_S24, ORF8_Y31*, ORF9b_S50*, ORF9b_V15, S_A222, S_D950*, S_G142*, S_K1191*, S_L212*, S_L5, S_N501, S_P681, S_R158*, S_R346*, S_S112*, S_T478, S_T732, S_T95, S_V1264*, S_V289

The XGBoost-SHAP analyses included six WHO variant features, Alpha, Delta, Epsilon, Gamma, Mu, and Omicron. Amino acid changes that were highly correlated with these, and consequently not included as independent features, were as follows:

Alpha: N_D3, N_S235, ORF1a_A1708, ORF1a_I2230, ORF1a_T1001, ORF8_Q27stop, ORF8_Y73, ORF8_R52, S_A570, S_D1118, S_S982, S_T716

Delta: M_I82, N_D63, N_D377, ORF1b_G662, ORF1b_P1000, ORF3a_S26, ORF7a_T120, ORF7a_V82, ORF9b_T60, S_L452, S_T19

© 2024 The MITRE Corporation, All Rights Reserved
Approved for Public Release. Distribution Unlimited. Public Release Case Number 24-0748.

Epsilon: ORF1b_D1183, S_S13, S_W152

Gamma: N_P80, ORF1a_K1795, ORF1a_S1188, ORF1b_E1264, ORF3a_S253, ORF8_E92, ORF9b_Q77, S_R190, S_T1027, S_T20, S_V1176

Mu: ORF1a_T1055, ORF1a_T1538, ORF1b_P1342, S_Y144

Omicron: E_T9, M_A63, M_D3, M_Q19, N_P13, ORF1a_A2710, ORF1a_I3758, ORF1a_K856, ORF1a_L2084, ORF1a_P3395, ORF1b_I1566, ORF9b_P10, S_D796, S_G339, S_G446, S_G496, S_A67, S_E484, S_H655, S_K417, S_L981, S_N440, S_N679, S_N764, S_N856, S_N969, S_Q493, S_Q498, S_Q954, S_S371, S_S373, S_S375, S_S477, S_T547, S_Y505, S_Y145

**GLMM Methods and Results**

To assess the overall ability of the fitted GLMMs to predict risk for hospitalization, we generated posterior predictive samples for each model fit (5000 predictions for each person). We then took the mean of 5000 predictions for each person, to arrive at an overall predicted probability of admission across predictive sampling replicates. We binned these predicted probabilities into 10 equally sized groups. For each bin, we calculated the actual probability of admission for the bin by referring to the true outcome for all persons in the bin. For each person in the bin, we simulated 500 binomial trials according to the person's predicted probability of admission and counted the number of successes in these trials. We then used these simulated counts to determine the simulated mean, 2.5%, and 97.5% quantiles for probability of admission across all persons in the group. Additionally, we examined the distribution of predicted probability of admission according to fitted models for the group of patients who were admitted compared to those who were not admitted.

Figure S5 shows a representative comparison of actual percent of persons admitted to simulated percent of persons admitted based on the fitted model, across 10 levels of model predicted risk. The figure shows overall reasonable agreement between simulations based on the fitted model and actual numbers across the 10 levels of risk. In all cases the actual percent admitted is within a 95% support interval defined by the model.

Additionally, we examined the distribution of predicted probability of admission according to fitted models for the group of patients who were admitted compared to those who were not admitted. Figure S6 shows a representative example of the distributions of predicted probability of admission according to the fitted model, comparing the group of persons who were actually admitted to those who were not. The figure demonstrates a shift toward higher predicted probability of admission among those persons who were actually admitted.

© 2024 The MITRE Corporation, All Rights Reserved
Approved for Public Release. Distribution Unlimited. Public Release Case Number 24-0748.

### Supplementary Tables and Figures

**Table S1**. Features used in disease risk score modeling. Each row is a feature used to build the disease risk score. Features that indicate conditions, e.g., condition_acquired_heart_disease_vs, were derived from diagnosis codes and value sets indicative of the corresponding feature. More information on these and other features included can be found in (23).

| Feature | Description / possible values |
|---|---|
| age_group | age, 10-year increments (e.g., [20,30), [30,40), etc.) |
| birthsex | male or female |
| condition_acquired_heart_disease_vs | present or absent |
| condition_acquired_immune_deficiency_syndrome_vs | present or absent |
| condition_alcohol_abuse_vs | present or absent |
| condition_arthropathies_vs | present or absent |
| condition_asthma_vs | present or absent |
| condition_cad_vs | present or absent |
| condition_cardiac_arrythmia_vs | present or absent |
| condition_cardiomyopathy_vs | present or absent |
| condition_cerebrovascular_disease_vs | present or absent |
| condition_chronic_blood_loss_anemia_vs | present or absent |
| condition_coagulopathy_vs | present or absent |
| condition_congenital_heart_disease_vs | present or absent |
| condition_copd_vs | present or absent |
| condition_cystic_fibrosis_vs | present or absent |
| condition_deficiency_anemias_vs | present or absent |
| condition_dementia_vs | present or absent |
| condition_depression_vs | present or absent |
| condition_diabetes_with_chronic_complications_vs | present or absent |
| condition_diabetes_without_chronic_complications_vs | present or absent |
| condition_down_syndrome_vs | present or absent |
| condition_drug_abuse_vs | present or absent |
| condition_heart_failure_vs | present or absent |
| condition_hypercoagulable_state_vs | present or absent |
| condition_hypertension_complicated_vs | present or absent |
| condition_hypertension_uncomplicated_vs | present or absent |

© 2024 The MITRE Corporation, All Rights Reserved
Approved for Public Release. Distribution Unlimited. Public Release Case Number 24-0748.

| | |
|---|---|
| condition_hypothyroidism_vs | present or absent |
| condition_interstitial_lung_disease_vs | present or absent |
| condition_leukemia_vs | present or absent |
| condition_liver_disease_mild_vs | present or absent |
| condition_liver_disease_moderate_to_severe_vs | present or absent |
| condition_lymphoma_vs | present or absent |
| condition_metastatic_cancer_vs | present or absent |
| condition_neurodevelopmental_disorders_vs | present or absent |
| condition_neurological_disorders_affecting_movement_vs | present or absent |
| condition_other_chronic_respiratory_disease_vs | present or absent |
| condition_other_immune_deficiency_vs | present or absent |
| condition_other_neurological_disorders_vs | present or absent |
| condition_other_thyroid_disorders_vs | present or absent |
| condition_pancreatitis_vs | present or absent |
| condition_paralysis_vs | present or absent |
| condition_peptic_ulcer_disease_vs | present or absent |
| condition_peripheral_vascular_disease_vs | present or absent |
| condition_psychoses_vs | present or absent |
| condition_pulmonary_circulation_disorders_vs | present or absent |
| condition_renal_failure_moderate_vs | present or absent |
| condition_renal_failure_severe_vs | present or absent |
| condition_seizures_and_epilepsy_vs | present or absent |
| condition_sickle_cell_disease_vs | present or absent |
| condition_solid_organ_or_blood_stem_cell_transplantation_vs | present or absent |
| condition_solid_tumor_without_metastasis_in_situ_vs | present or absent |
| condition_solid_tumor_without_metastasis_malignant_vs | present or absent |
| condition_thalassemia_vs | present or absent |
| condition_valvular_disease_vs | present or absent |
| condition_weight_loss_vs | present or absent |
| ethnicity | Hispanic/Latino or not |
| health_system | variable representing which of 4 health systems |
| immunosuppressant_prev90days | present or absent |
| marital_status | married, unmarried, divorced, widowed |
| insurance_category | private, medicare, medicaid, self-pay, military, other |

© 2024 The MITRE Corporation, All Rights Reserved
Approved for Public Release. Distribution Unlimited. Public Release Case Number 24-0748.

| | |
|---|---|
| obese | present or absent |
| out_of_state | whether or not patient lives in same state as health system |
| pregnant | true or false at time of COVID-19 diagnosis |
| race | White, Black/African American, Asian, Native Hawaiian/Other Pacific Islander, American Indian/Alaska Native, Other |
| smoke_status | true or false at time of COVID-19 diagnosis |
| total_visits | count of visits to health system in 24 months prior to COVID19 diagnosis |
| zip3_pop_density | population density of patient's 3-digit zip code |
| zip3_adi | area deprivation index of patient's 3-digit zip code |

© 2024 The MITRE Corporation, All Rights Reserved
Approved for Public Release. Distribution Unlimited. Public Release Case Number 24-0748.

**Table S2**. Mapping from lineage designations to WHO variants.

| WHO variant | Lineage | Number of samples |
|---|---|---|
| Alpha | B.1.1.7 | 1090 |
| Alpha | Q.3 | 22 |
| Delta | AY.1 | 4 |
| Delta | AY.100 | 685 |
| Delta | AY.103 | 1352 |
| Delta | AY.105 | 5 |
| Delta | AY.107 | 25 |
| Delta | AY.109 | 1 |
| Delta | AY.110 | 8 |
| Delta | AY.113 | 37 |
| Delta | AY.114 | 15 |
| Delta | AY.116 | 1 |
| Delta | AY.116.1 | 16 |
| Delta | AY.117 | 42 |
| Delta | AY.118 | 47 |
| Delta | AY.119 | 65 |
| Delta | AY.119.2 | 8 |
| Delta | AY.120 | 40 |
| Delta | AY.120.1 | 11 |
| Delta | AY.121 | 2 |
| Delta | AY.122 | 42 |
| Delta | AY.125 | 1 |
| Delta | AY.127 | 3 |
| Delta | AY.129 | 16 |
| Delta | AY.13 | 22 |
| Delta | AY.14 | 32 |
| Delta | AY.16 | 6 |
| Delta | AY.2 | 11 |
| Delta | AY.20 | 65 |
| Delta | AY.25 | 1015 |
| Delta | AY.25.1 | 166 |
| Delta | AY.25.3 | 1 |
| Delta | AY.26 | 163 |
| Delta | AY.3 | 665 |
| Delta | AY.3.1 | 61 |
| Delta | AY.3.3 | 1 |
| Delta | AY.34 | 2 |

© 2024 The MITRE Corporation, All Rights Reserved
Approved for Public Release. Distribution Unlimited. Public Release Case Number 24-0748.

| | | |
|---|---|---|
| Delta | AY.34.1 | 4 |
| Delta | AY.35 | 10 |
| Delta | AY.36 | 4 |
| Delta | AY.37 | 9 |
| Delta | AY.39 | 174 |
| Delta | AY.39.1 | 9 |
| Delta | AY.4 | 30 |
| Delta | AY.42 | 2 |
| Delta | AY.43 | 7 |
| Delta | AY.44 | 1922 |
| Delta | AY.46 | 6 |
| Delta | AY.46.1 | 1 |
| Delta | AY.46.4 | 40 |
| Delta | AY.47 | 218 |
| Delta | AY.48 | 14 |
| Delta | AY.49 | 1 |
| Delta | AY.5 | 1 |
| Delta | AY.5.3 | 2 |
| Delta | AY.52 | 9 |
| Delta | AY.54 | 172 |
| Delta | AY.62 | 9 |
| Delta | AY.64 | 12 |
| Delta | AY.67 | 7 |
| Delta | AY.7.1 | 1 |
| Delta | AY.74 | 4 |
| Delta | AY.75 | 83 |
| Delta | AY.77 | 5 |
| Delta | AY.81 | 29 |
| Delta | AY.83 | 4 |
| Delta | AY.86 | 2 |
| Delta | AY.88 | 1 |
| Delta | AY.9.2 | 7 |
| Delta | AY.98.1 | 7 |
| Delta | B.1.617.2 | 370 |
| earlyVar | B.1 | 57 |
| earlyVar | B.1.1 | 8 |
| earlyVar | B.1.1.186 | 1 |
| earlyVar | B.1.1.192 | 1 |
| earlyVar | B.1.1.222 | 61 |
| earlyVar | B.1.1.239 | 1 |

© 2024 The MITRE Corporation, All Rights Reserved
Approved for Public Release. Distribution Unlimited. Public Release Case Number 24-0748.

| | | |
|---|---|---|
| earlyVar | B.1.1.251 | 1 |
| earlyVar | B.1.1.318 | 7 |
| earlyVar | B.1.1.348 | 1 |
| earlyVar | B.1.1.416 | 10 |
| earlyVar | B.1.1.432 | 3 |
| earlyVar | B.1.1.519 | 80 |
| earlyVar | B.1.111 | 2 |
| earlyVar | B.1.126 | 1 |
| earlyVar | B.1.2 | 339 |
| earlyVar | B.1.232 | 2 |
| earlyVar | B.1.234 | 11 |
| earlyVar | B.1.239 | 4 |
| earlyVar | B.1.240 | 1 |
| earlyVar | B.1.241 | 2 |
| earlyVar | B.1.243 | 13 |
| earlyVar | B.1.258.23 | 11 |
| earlyVar | B.1.265 | 2 |
| earlyVar | B.1.349 | 1 |
| earlyVar | B.1.351 | 9 |
| earlyVar | B.1.36.7 | 1 |
| earlyVar | B.1.369 | 2 |
| earlyVar | B.1.375 | 1 |
| earlyVar | B.1.396 | 2 |
| earlyVar | B.1.400 | 16 |
| earlyVar | B.1.426 | 1 |
| earlyVar | B.1.433 | 2 |
| earlyVar | B.1.525 | 2 |
| earlyVar | B.1.526 | 21 |
| earlyVar | B.1.544 | 1 |
| earlyVar | B.1.551 | 3 |
| earlyVar | B.1.561 | 19 |
| earlyVar | B.1.565 | 1 |
| earlyVar | B.1.568 | 1 |
| earlyVar | B.1.575 | 11 |
| earlyVar | B.1.577 | 1 |
| earlyVar | B.1.582 | 4 |
| earlyVar | B.1.587 | 2 |
| earlyVar | B.1.595 | 9 |
| earlyVar | B.1.596 | 2 |
| earlyVar | B.1.599 | 1 |

© 2024 The MITRE Corporation, All Rights Reserved
Approved for Public Release. Distribution Unlimited. Public Release Case Number 24-0748.

| | | |
|---|---|---|
| earlyVar | B.1.609 | 6 |
| earlyVar | B.1.617.1 | 1 |
| earlyVar | B.1.627 | 1 |
| earlyVar | B.1.631 | 1 |
| earlyVar | B.1.634 | 1 |
| earlyVar | B.1.637 | 29 |
| Epsilon | B.1.427 | 65 |
| Epsilon | B.1.429 | 237 |
| Gamma | P.1 | 53 |
| Gamma | P.1.10 | 1 |
| Gamma | P.1.14 | 2 |
| Gamma | P.1.15 | 1 |
| Gamma | P.1.17 | 3 |
| Mu | B.1.621 | 69 |
| Mu | B.1.621.1 | 1 |
| Omicron | BA.1 | 268 |
| Omicron | BA.1.1 | 988 |
| Omicron | BA.1.1.10 | 1 |
| Omicron | BA.1.1.12 | 1 |
| Omicron | BA.1.1.14 | 15 |
| Omicron | BA.1.1.18 | 76 |
| Omicron | BA.1.1.2 | 14 |
| Omicron | BA.1.1.4 | 1 |
| Omicron | BA.1.1.8 | 1 |
| Omicron | BA.1.13 | 1 |
| Omicron | BA.1.14 | 2 |
| Omicron | BA.1.15 | 743 |
| Omicron | BA.1.15.1 | 3 |
| Omicron | BA.1.15.2 | 7 |
| Omicron | BA.1.17 | 14 |
| Omicron | BA.1.17.2 | 12 |
| Omicron | BA.1.18 | 79 |
| Omicron | BA.1.19 | 1 |
| Omicron | BA.1.20 | 156 |
| Omicron | BA.1.21 | 1 |
| Omicron | BA.2 | 2 |
| Omicron | BA.2.2.1 | 3 |
| Omicron | BA.2.3 | 1 |
| other | A.2.5 | 1 |
| other | AZ.3 | 5 |

© 2024 The MITRE Corporation, All Rights Reserved
Approved for Public Release. Distribution Unlimited. Public Release Case Number 24-0748.

| | | |
|---|---|---|
| other | C.37 | 4 |
| other | P.2 | 8 |
| other | R.1 | 3 |

© 2024 The MITRE Corporation, All Rights Reserved
Approved for Public Release. Distribution Unlimited. Public Release Case Number 24-0748.

**Table S3**. Hyperparameter grid for XGBoost models. Grid search was performed using `sklearn.model_selection.RandomizedSearchCV`. The hyperparameter `scale_pos_weight` was set to a constant value to reweight the data, due to the relatively rare outcome of hospitalization.

| Hyperparameter | Values evaluated |
|---|---|
| n_estimators | [100, 500, 1000, None] |
| max_depth | [3, 5, 6, 8, 10, 15, 20] |
| learning_rate | [0.05, 0.10, 0.20, 0.30] |
| min_child_weight | [0.5, 1, 3, 5, 7] |
| gamma | [0.0, 0.1, 0.2, 0.3, 0.4] |
| colsample_bytree | [0.3, 0.5, 0.7, 1.0] |
| scale_pos_weight | [24.5] |

© 2024 The MITRE Corporation, All Rights Reserved
Approved for Public Release. Distribution Unlimited. Public Release Case Number 24-0748.

**Table S4**. Findings with alternative priors for GLMMs. As discussed in Methods, GLMM parameter estimates were obtained using Bayesian techniques. We repeated the same analyses with a number of different prior distributions. The lineage intercept estimate priors were normally distributed, centered on the base admission rate, with a scale parameter $\sigma_0$. The amino acid variant effect priors were Laplace distributed, centered on 0, with a shared scale parameter $\sigma_1$. For the estimation of effects associated with amino acid changes, the value of $\sigma_0$ was set at 1, and $\sigma_1$ was varied as 0.0005, 0.001, 0.01, and 0.1. This table shows, for each combination of $\sigma_0$ and $\sigma_1$, the amino acid changes that were highlighted as potentially important. Essentially the same set of amino acid changes was highlighted with all priors. The table shows $\sigma_0$ and $\sigma_1$ (columns labeled sigma_0 and sigma_1), the lineage within which the effect of the amino acid change is estimated, the frequency of the amino acid change in the lineage and the count of samples in the lineage that have the change, and the z-score and Bayes factor of the posterior distribution.

| Sigma_0 prior | Sigma_1 prior | Amino acid change | Lineage | Amino acid change frequency | Number of samples | Z-score | Bayes factor |
|---|---|---|---|---|---|---|---|
| 1 | 0.0005 | N_M234 | B.1.429 | 0.388 | 92 | -1.55 | 6.13 |
| 1 | 0.0005 | ORF1a_R4179 | AY.25 | 0.312 | 317 | 1.48 | 4.48 |
| 1 | 0.0005 | ORF1b_P1975 | B.1.1.7 | 0.176 | 192 | 1.04 | 3.13 |
| 1 | 0.0005 | ORF3a_P104 | AY.103 | 0.083 | 112 | -1.10 | 3.13 |
| 1 | 0.0005 | ORF8_Y31 | AY.100 | 0.591 | 405 | -1.15 | 3.84 |
| 1 | 0.0005 | S_D950 | AY.39 | 0.305 | 53 | -1.05 | 3.25 |
| 1 | 0.0005 | S_G142 | AY.100 | 0.632 | 433 | -1.10 | 3.44 |
| 1 | 0.0005 | S_G142 | AY.3 | 0.284 | 189 | -1.28 | 4.39 |
| 1 | 0.0005 | S_L212 | BA.1 | 0.653 | 175 | 1.29 | 3.67 |
| 1 | 0.0005 | S_S112 | AY.25 | 0.470 | 477 | 1.01 | 3.12 |
| 1 | 0.001 | N_M234 | B.1.429 | 0.388 | 92 | -1.55 | 6.13 |
| 1 | 0.001 | ORF1a_R4179 | AY.25 | 0.312 | 317 | 1.48 | 4.48 |
| 1 | 0.001 | ORF1b_P1975 | B.1.1.7 | 0.176 | 192 | 1.04 | 3.13 |
| 1 | 0.001 | ORF3a_P104 | AY.103 | 0.083 | 112 | -1.10 | 3.13 |
| 1 | 0.001 | ORF8_Y31 | AY.100 | 0.591 | 405 | -1.15 | 3.84 |
| 1 | 0.001 | S_D950 | AY.39 | 0.305 | 53 | -1.05 | 3.25 |
| 1 | 0.001 | S_G142 | AY.100 | 0.632 | 433 | -1.10 | 3.44 |
| 1 | 0.001 | S_G142 | AY.3 | 0.284 | 189 | -1.28 | 4.39 |
| 1 | 0.001 | S_L212 | BA.1 | 0.653 | 175 | 1.29 | 3.67 |
| 1 | 0.001 | S_S112 | AY.25 | 0.470 | 477 | 1.01 | 3.12 |
| 1 | 0.01 | N_M234 | B.1.429 | 0.388 | 92 | -1.55 | 6.13 |
| 1 | 0.01 | ORF1a_R4179 | AY.25 | 0.312 | 317 | 1.48 | 4.48 |
| 1 | 0.01 | ORF1b_P1975 | B.1.1.7 | 0.176 | 192 | 1.04 | 3.13 |
| 1 | 0.01 | ORF3a_P104 | AY.103 | 0.083 | 112 | -1.10 | 3.13 |

© 2024 The MITRE Corporation, All Rights Reserved
Approved for Public Release. Distribution Unlimited. Public Release Case Number 24-0748.

| | | | | | | | |
|---|---|---|---|---|---|---|---|
| 1 | 0.01 | ORF8_Y31 | AY.100 | 0.591 | 405 | -1.15 | 3.84 |
| 1 | 0.01 | S_D950 | AY.39 | 0.305 | 53 | -1.05 | 3.25 |
| 1 | 0.01 | S_G142 | AY.100 | 0.632 | 433 | -1.10 | 3.44 |
| 1 | 0.01 | S_G142 | AY.3 | 0.284 | 189 | -1.28 | 4.39 |
| 1 | 0.01 | S_L212 | BA.1 | 0.653 | 175 | 1.29 | 3.67 |
| 1 | 0.01 | S_S112 | AY.25 | 0.470 | 477 | 1.01 | 3.12 |
| 1 | 0.1 | N_M234 | B.1.429 | 0.388 | 92 | -1.55 | 6.13 |
| 1 | 0.1 | ORF1a_R4179 | AY.25 | 0.312 | 317 | 1.48 | 4.48 |
| 1 | 0.1 | ORF1b_P1975 | B.1.1.7 | 0.176 | 192 | 1.04 | 3.13 |
| 1 | 0.1 | ORF3a_P104 | AY.103 | 0.083 | 112 | -1.10 | 3.13 |
| 1 | 0.1 | ORF8_Y31 | AY.100 | 0.591 | 405 | -1.15 | 3.84 |
| 1 | 0.1 | S_D950 | AY.39 | 0.305 | 53 | -1.05 | 3.25 |
| 1 | 0.1 | S_G142 | AY.100 | 0.632 | 433 | -1.10 | 3.44 |
| 1 | 0.1 | S_G142 | AY.3 | 0.284 | 189 | -1.28 | 4.39 |
| 1 | 0.1 | S_L212 | BA.1 | 0.653 | 175 | 1.29 | 3.67 |
| 1 | 0.1 | S_S112 | AY.25 | 0.470 | 477 | 1.01 | 3.12 |

© 2024 The MITRE Corporation, All Rights Reserved
Approved for Public Release. Distribution Unlimited. Public Release Case Number 24-0748.

**Table S5**. Stability of parameter estimates across 28 runs of the GLMM model, one run for each amino acid change. Shown are average value of the mean parameter estimates and standard error of the mean across all runs of GLMMs. Each of the parameters shown appears across all 28 runs of the GLMM model. The standard errors are relatively small, generally on the order of 0.02-0.03, indicating that estimates for $\alpha_1$, $\alpha_2$, $\alpha_3$, $\sigma_0$, $\sigma_1$, and the lineage intercept estimates (shown for the 30 most common lineages, e.g., AY.100, AY.103, etc.) are relatively stable across runs for different amino acid changes.

| Feature | Mean | Std | Min | 25% | 50% | 75% | Max |
|---|---|---|---|---|---|---|---|
| alpha_1 | -1.11 | 0.01 | -1.13 | -1.12 | -1.11 | -1.11 | -1.09 |
| alpha_2 | 3.67 | 0.02 | 3.64 | 3.66 | 3.66 | 3.68 | 3.72 |
| alpha_3 | -1.04 | 0.01 | -1.05 | -1.04 | -1.04 | -1.03 | -1.02 |
| sigma_0 | 0.40 | 0.00 | 0.40 | 0.40 | 0.40 | 0.40 | 0.40 |
| sigma_1 | 0.35 | 0.01 | 0.33 | 0.34 | 0.35 | 0.36 | 0.38 |
| AY.100 | -3.02 | 0.04 | -3.17 | -3.02 | -3.01 | -3.00 | -2.98 |
| AY.103 | -2.95 | 0.01 | -2.97 | -2.95 | -2.95 | -2.94 | -2.90 |
| AY.119 | -3.46 | 0.02 | -3.51 | -3.47 | -3.46 | -3.45 | -3.42 |
| AY.20 | -2.89 | 0.04 | -2.95 | -2.92 | -2.90 | -2.88 | -2.81 |
| AY.25 | -3.07 | 0.02 | -3.10 | -3.08 | -3.06 | -3.06 | -3.01 |
| AY.25.1 | -2.95 | 0.03 | -3.00 | -2.97 | -2.96 | -2.93 | -2.89 |
| AY.26 | -3.19 | 0.03 | -3.24 | -3.21 | -3.18 | -3.17 | -3.13 |
| AY.3 | -3.02 | 0.02 | -3.05 | -3.03 | -3.02 | -3.01 | -2.98 |
| AY.3.1 | -3.25 | 0.04 | -3.31 | -3.27 | -3.25 | -3.22 | -3.17 |
| AY.39 | -3.29 | 0.02 | -3.31 | -3.30 | -3.29 | -3.27 | -3.23 |
| AY.44 | -2.82 | 0.03 | -2.87 | -2.84 | -2.83 | -2.82 | -2.74 |
| AY.47 | -3.07 | 0.03 | -3.12 | -3.09 | -3.06 | -3.05 | -3.01 |
| AY.54 | -2.68 | 0.03 | -2.74 | -2.70 | -2.68 | -2.66 | -2.64 |
| AY.75 | -3.25 | 0.04 | -3.32 | -3.28 | -3.26 | -3.24 | -3.17 |
| B.1 | -2.94 | 0.03 | -2.99 | -2.96 | -2.93 | -2.91 | -2.86 |
| B.1.1.222 | -3.12 | 0.03 | -3.18 | -3.14 | -3.12 | -3.10 | -3.07 |
| B.1.1.519 | -2.92 | 0.03 | -2.97 | -2.94 | -2.92 | -2.90 | -2.88 |
| B.1.1.7 | -2.71 | 0.03 | -2.84 | -2.71 | -2.71 | -2.69 | -2.69 |
| B.1.2 | -3.43 | 0.01 | -3.47 | -3.44 | -3.44 | -3.42 | -3.42 |
| B.1.427 | -3.43 | 0.03 | -3.49 | -3.45 | -3.43 | -3.41 | -3.36 |
| B.1.429 | -3.45 | 0.02 | -3.48 | -3.47 | -3.45 | -3.44 | -3.42 |
| B.1.617.2 | -3.42 | 0.03 | -3.47 | -3.44 | -3.42 | -3.41 | -3.34 |
| B.1.621 | -2.65 | 0.03 | -2.70 | -2.67 | -2.66 | -2.62 | -2.58 |
| BA.1 | -3.63 | 0.02 | -3.70 | -3.64 | -3.63 | -3.62 | -3.56 |
| BA.1.1 | -3.47 | 0.02 | -3.50 | -3.48 | -3.46 | -3.45 | -3.43 |
| BA.1.1.18 | -3.16 | 0.03 | -3.21 | -3.18 | -3.16 | -3.13 | -3.10 |
| BA.1.15 | -3.35 | 0.01 | -3.37 | -3.36 | -3.35 | -3.34 | -3.32 |

© 2024 The MITRE Corporation, All Rights Reserved
Approved for Public Release. Distribution Unlimited. Public Release Case Number 24-0748.

| | | | | | | | |
|---|---|---|---|---|---|---|---|
| BA.1.18 | -3.43 | 0.02 | -3.47 | -3.44 | -3.42 | -3.41 | -3.38 |
| BA.1.20 | -3.53 | 0.03 | -3.58 | -3.55 | -3.53 | -3.51 | -3.49 |
| P.1 | -3.11 | 0.04 | -3.19 | -3.13 | -3.11 | -3.09 | -3.01 |

© 2024 The MITRE Corporation, All Rights Reserved
Approved for Public Release. Distribution Unlimited. Public Release Case Number 24-0748.

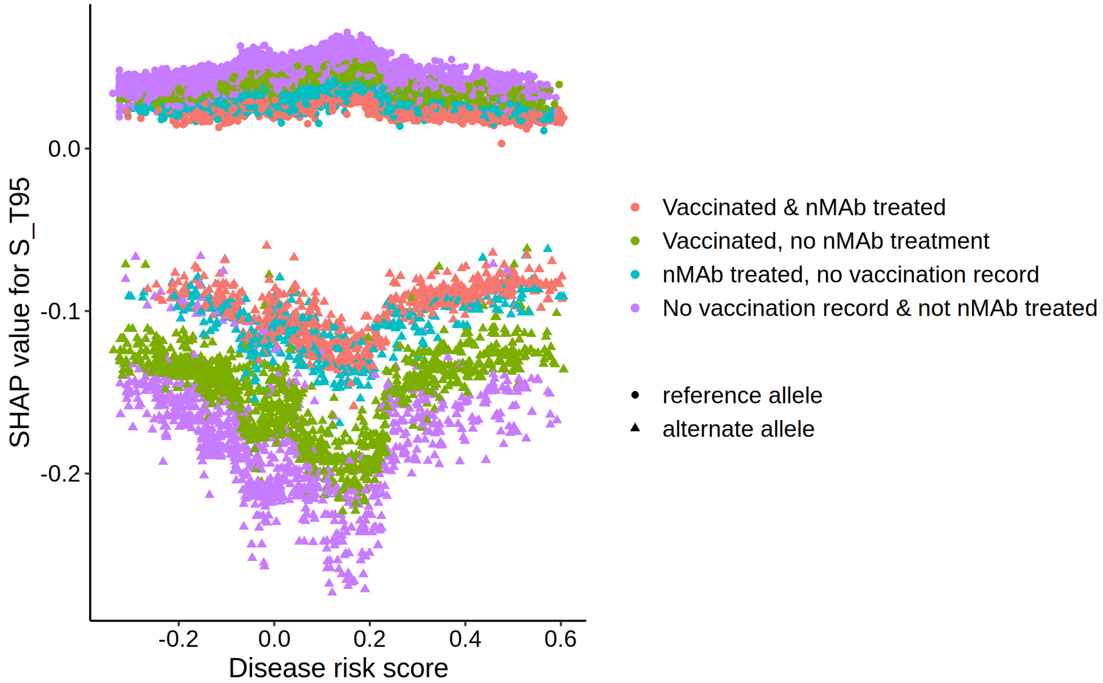


**Figure S1**. Median SHAP values for S_T95 for all samples by S_T95 alternate/reference allele, disease risk score, COVID-19 vaccination, and nMAb treatment status.

© 2024 The MITRE Corporation, All Rights Reserved
Approved for Public Release. Distribution Unlimited. Public Release Case Number 24-0748.

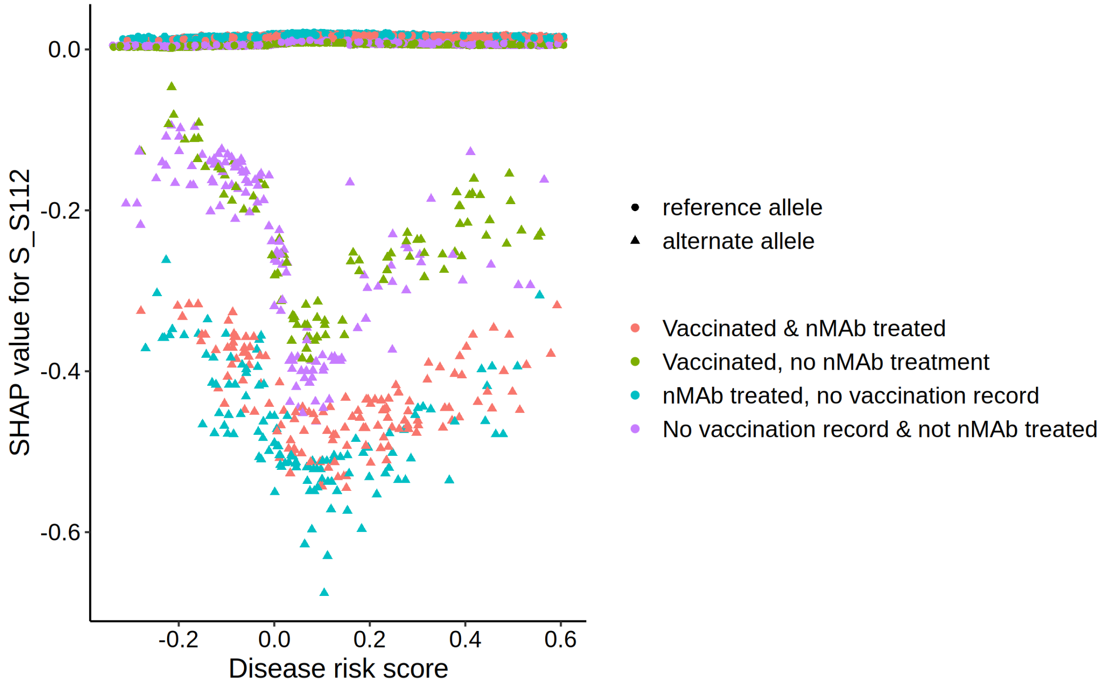


**Figure S2**. Median SHAP values for S_S112 for all samples by S_S112 alternate/reference allele, disease risk score, COVID-19 vaccination, and nMAb treatment status. There were 483 samples with an alternate substitution at S_S112. All occurred within Delta and had the mutation S_S112L.

© 2024 The MITRE Corporation, All Rights Reserved
Approved for Public Release. Distribution Unlimited. Public Release Case Number 24-0748.

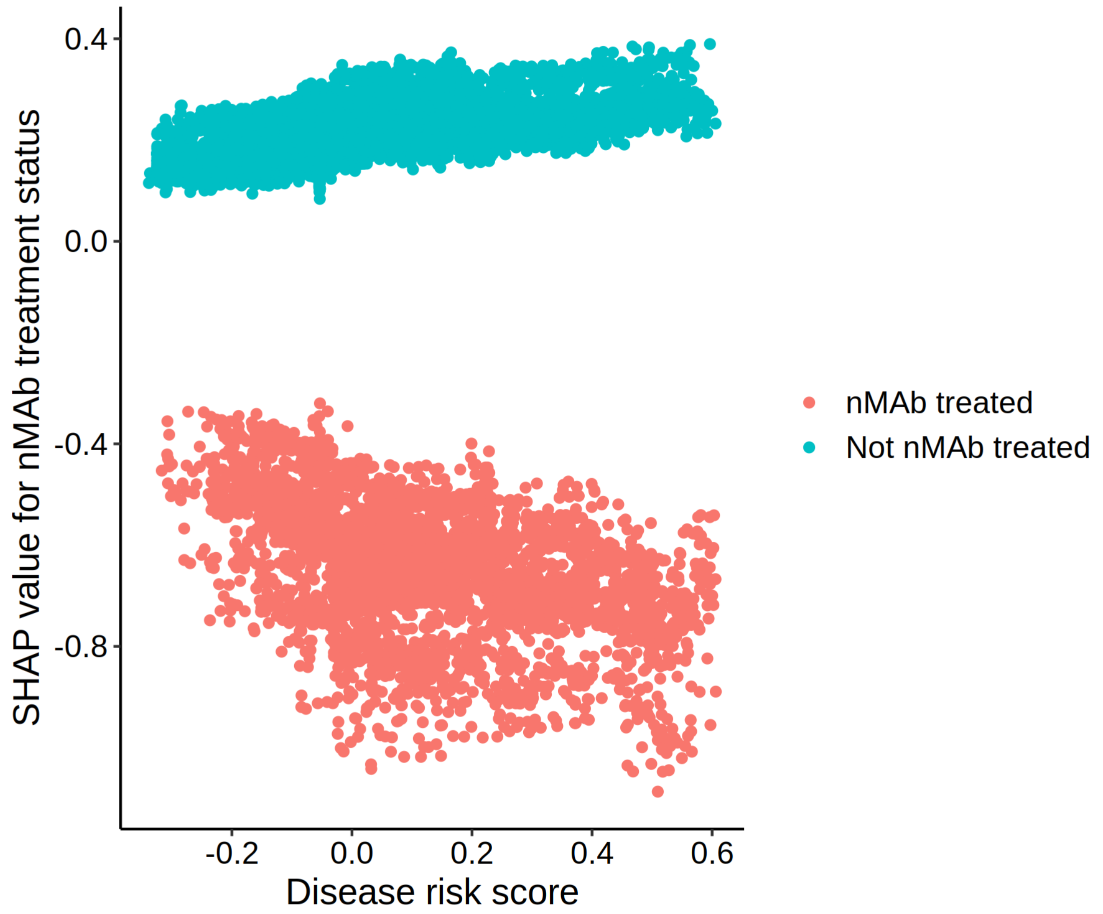


**Figure S3**. Median SHAP values for nMAb treatment for all samples by nMAb treatment status and disease risk score.

© 2024 The MITRE Corporation, All Rights Reserved
Approved for Public Release. Distribution Unlimited. Public Release Case Number 24-0748.

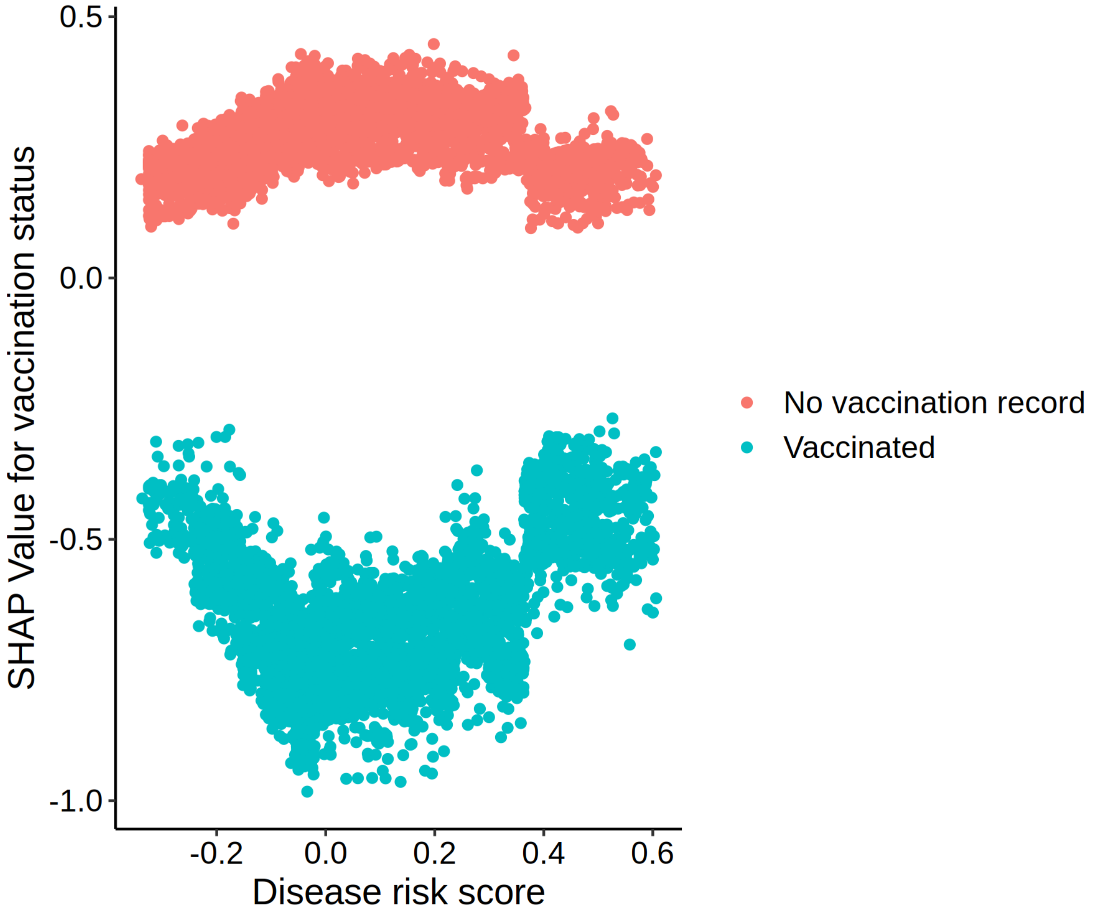


**Figure S4**. Median SHAP values for COVID-19 vaccination for all samples by vaccination status and disease risk score.

© 2024 The MITRE Corporation, All Rights Reserved
Approved for Public Release. Distribution Unlimited. Public Release Case Number 24-0748.

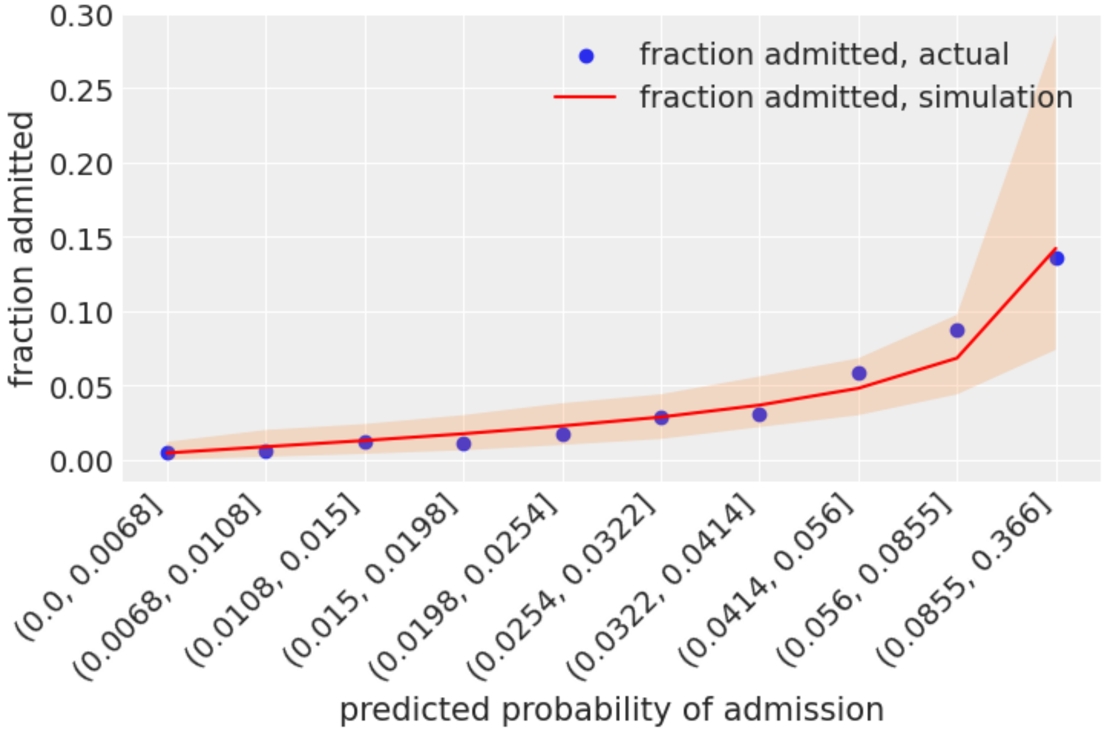


**Figure S5**. Representative comparison of actual versus simulated percent persons admitted based on the fitted model, across 10 levels, or bins of model predicted risk. Actual percent admitted in each bin is shown as a dot. Mean expected percent admitted in each bin, according to the fitted model, is shown as a red line. The shaded region shows a 95% support interval for simulated percent admitted.

© 2024 The MITRE Corporation, All Rights Reserved
Approved for Public Release. Distribution Unlimited. Public Release Case Number 24-0748.

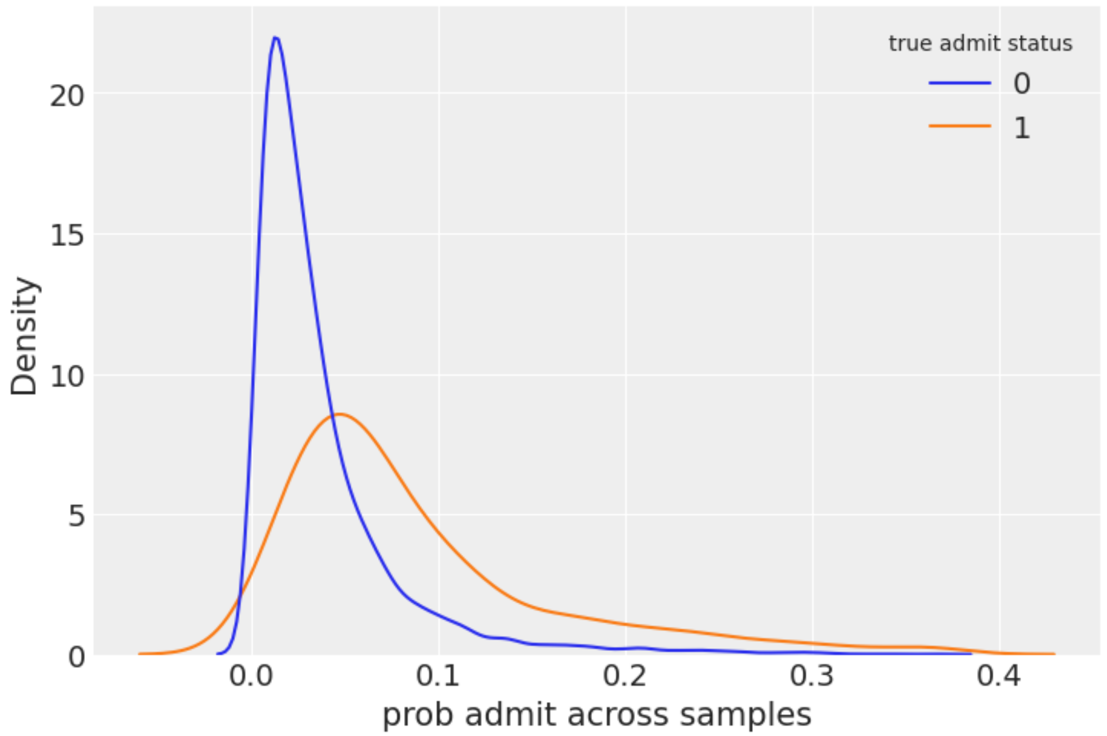


**Figure S6**. Representative example of predicted probability of admission distributions for those patients who were actually admitted (orange) compared to those who were not admitted (blue).

© 2024 The MITRE Corporation, All Rights Reserved
Approved for Public Release. Distribution Unlimited. Public Release Case Number 24-0748.
